# Common and rare genetic variation intersects with ancestry to influence human skin and plasma carotenoid concentrations

**DOI:** 10.1101/2024.12.20.24319465

**Authors:** Yixing Han, Savannah Mwesigwa, Qiang Wu, Melissa N. Laska, Stephanie B. Jilcott Pitts, Nancy E. Moran, Neil A. Hanchard

## Abstract

Carotenoids are dietary bioactive compounds with health effects that are biomarkers of fruit and vegetable intake. Here, we examine genetic associations with plasma and skin carotenoid concentrations in two rigorously phenotyped human cohorts (n=317). Analysis of genome-wide SNPs revealed heritability to vary by genetic ancestry (h²=0.08–0.44) with ten SNPs at four loci reaching genome-wide significance (P<5E-08) in multivariate models, including at *RAPGEF1* (rs3765544, P=8.86E-10, beta=0.75) with α-carotene, and near *IGSF11* (rs80316816, P=6.25E-10, beta=0.74), with cryptoxanthin; these were replicated in the second cohort (n=110). Multiple SNPs near *IGSF11* demonstrated genotype-dependent dietary effects on plasma cryptoxanthin. Deep sequencing of 35 candidate genes revealed associations between the *PKD1L2*-*BCO1* locus and plasma β-carotene (Padj=0.04, beta=-1.3 to -0.3), and rare, ancestry-restricted, damaging variants in *CETP* (rs2303790) and *APOA1* (rs756535387) in individuals with high skin carotenoids. Our findings implicate novel loci in carotenoid disposition and indicate the importance of including cohorts of diverse genetic ancestry.

## INTRODUCTION

Carotenoids are a diverse group of natural pigments produced by plants, fungi, algae, and photosynthetic bacteria. While there are over 1000 identified carotenoid species in nature^1^, six species (α-carotene, β-carotene, cryptoxanthin, lycopene, lutein, and zeaxanthin) constitute more than 95% of total human blood carotenoids^2^. Humans cannot synthesize carotenoids endogenously, thus primarily acquire carotenoids from dietary fruits and vegetables (FV), mostly in the form of β-carotene, lycopene, and lutein/zeaxanthin^3,4^. Carotenoids are absorbed, metabolized, and distributed throughout the blood, skin, and other tissues in a manner similar to dietary lipids^4^. Carotenoid activities depend on their chemical properties, which can be pro-vitamin A, nuclear receptor signaling, light filtering, or antioxidant/anti-inflammatory^3,4^. Because carotenoids are absorbed, retained in the body for a moderate amount of time, and are detectable by spectroscopy, carotenoid concentrations, measured in plasma and more recently non-invasively measured in the skin, have been proffered as biomarkers for dietary fruit and vegetable intake assessment in adults and children^5,6^.

Epidemiologic, clinical, and preclinical studies also indicate that carotenoids are associated with protection from many chronic diseases, including cancers, cardiovascular disease, and macular degeneration^4^. Their importance lies in their ability to modulate intracellular signaling, offering antioxidant, antiapoptotic, and anti-inflammatory properties that protect cells from oxidative stress, UV damage, and support functions like vision and immune response^3^.

While carotenoid plasma and tissue concentrations are primarily a function of dietary intake, there is still substantive inter-individual variation in plasma and tissue carotenoid concentrations. Age, body mass index (BMI), and smoking have all been implicated as environmental contributors to this inter-individual variation. Genetic variation is known to also contribute to this variation^4,7–10^. For instance, in Mexican American children, the heritability of plasma carotenoid concentrations has been quantified, with α-carotene demonstrating a heritability (h²) of 0.81 (P = 6.7 × 10E-11) and β-carotene exhibiting an even higher heritability of 0.90 (P = 3.5 × 10E-15)^8^. However, a detailed understanding of how genetic variation influences interindividual remains unclear. A handful of early epidemiologic and clinical studies found associations between common SNPs in select lipid and carotenoid metabolism genes and blood concentrations of specific carotenoid species^11–15^. An early genome-wide association study (GWAS) in European populations identified an association with genetic variation near *β- carotene 15,15-dioxygenase (BCO1)*^12^, the key enzyme responsible for central cleavage of provitamin A carotenoids to yield vitamin A^16^. At the tissue-specific level even less is known about the impact of genetic variation on carotenoid levels; carotenoids in the macula of the eye have been associated with SNPs in *BCO1, BCO2, NPC1L1, ABCG8*, and *FADS2*^17–19^, and in small studies, carotenoids in prostate and skin have been associated with SNPs in the same genes^9,20^, albeit without subsequent replication of results.

To date, a broadly comprehensive understanding of how human genetic variation influences plasma and tissue carotenoid concentrations, particularly bioactive carotenoid species, remains elusive. This knowledge gap is strikingly evident for populations with non-European ancestral backgrounds, as most prior studies have focused on individuals of European descent. For example, early studies linking variants in *BCMO1* (aka *BCO1*) to plasma β-carotene levels^12^ and variants in *RBP4 (*retinol-binding protein 4) to circulating retinol (a carotenoid metabolite) levels^21^ were both conducted in homogenous Eurocentric cohorts. Additionally, *SCARB1* (a key receptor for carotenoid uptake) has been associated with plasma lycopene levels in multiethnic populations under certain conditions such as in postmenopausal women, though the effect sizes vary across groups^22^. The lack of unbiased genome-wide analyses, particularly in ancestrally diverse cohorts, restricts the generalizability of carotenoid genetics findings, limiting their applicability in both population and precision nutrition strategies^23–25^. Conducting comprehensive studies in diverse populations is thus essential going forward, as efforts to implement precision medicine and nutrigenomic initiatives, and identify biomarkers that can be used across population groups, intensify.

Here, we leverage detailed phenotypic data (including age, sex, BMI, self-reported race/ethnicity, and food intake), and rigorous plasma and skin carotenoid assessments from two ancestrally and geographically diverse US cohorts to conduct both genome-wide and targeted (sequence-based) association studies of plasma carotenoid species and skin carotenoid levels. We identify novel population common- and rare-genetic variants associated with steady-state and diet-responsive carotenoid levels. Further, we reveal ancestry-specific differences in heritability and genetic association. Using data derived from a controlled dietary intake study, we also uncover gene-by-dosage interactions at associated loci. Collectively, our findings highlight previously unrecognized genetic heterogeneity in human carotenoid metabolism, providing a foundation for advancing precision nutrition and understanding global health and nutrigenetics.

## RESULTS

Genetic studies were conducted in two extensively characterized, previously described cohorts^26,27^ (**Methods**). The primary discovery cohort comprised 213 individuals (207 after QC and familial relatedness check) from four self-reported United States (US) racial and ethnic groups (Asian, Hispanic, Non-Hispanic Black, and Non-Hispanic White) (**Supplementary Table S1**). Participants were recruited from two sites in the US, and had extensive clinical, lifestyle, and demographic phenotype data documented alongside cross-sectional plasma carotenoid species concentrations (measured by HPLC-photodiode array detection) and aggregate skin carotenoids (skin carotenoid score - measured by non-invasive pressure mediated reflection spectroscopy). These individuals were genotyped using the H3Africa genotyping microarray (Illumina, CA, USA), designed to be used in genetically diverse populations. To cover candidate loci that were not well-represented on the genotyping array, short-read capture-based sequencing was performed across 35 genomic loci, which are reported to influence carotenoid concentrations (**Methods, Supplementary Table S7**).

The secondary (intervention) cohort consisted of 162 individuals (**Supplementary Table S2**), of whom 110 unique participants were not included in the primary cohort. Participants were recruited from three sites, two of which were the same as the primary cohort, as part of a dietary carotenoid intervention study. This study collected identical phenotypic data within a longitudinal, randomized carotenoid-rich juice dose-response study. Data were collected at baseline, 3-, and 6-weeks post-intervention for three mixed carotenoid doses (low/control (0 mg/d), moderate (4 mg/d), and high (8 mg/d)). This cohort underwent the same genomic interrogation using the same platform and panel as the discovery cohort (**Figure 1**) (**Methods and Supplementary Methods**). For genetic analyses, self-reported race/ethnicity was refined through Multidimensional Scaling (MDS) of genomic information, aligning participants with human ancestral populations based on the 1000 Genomes Project data (**Figure 2A**).

**Figure 1.**
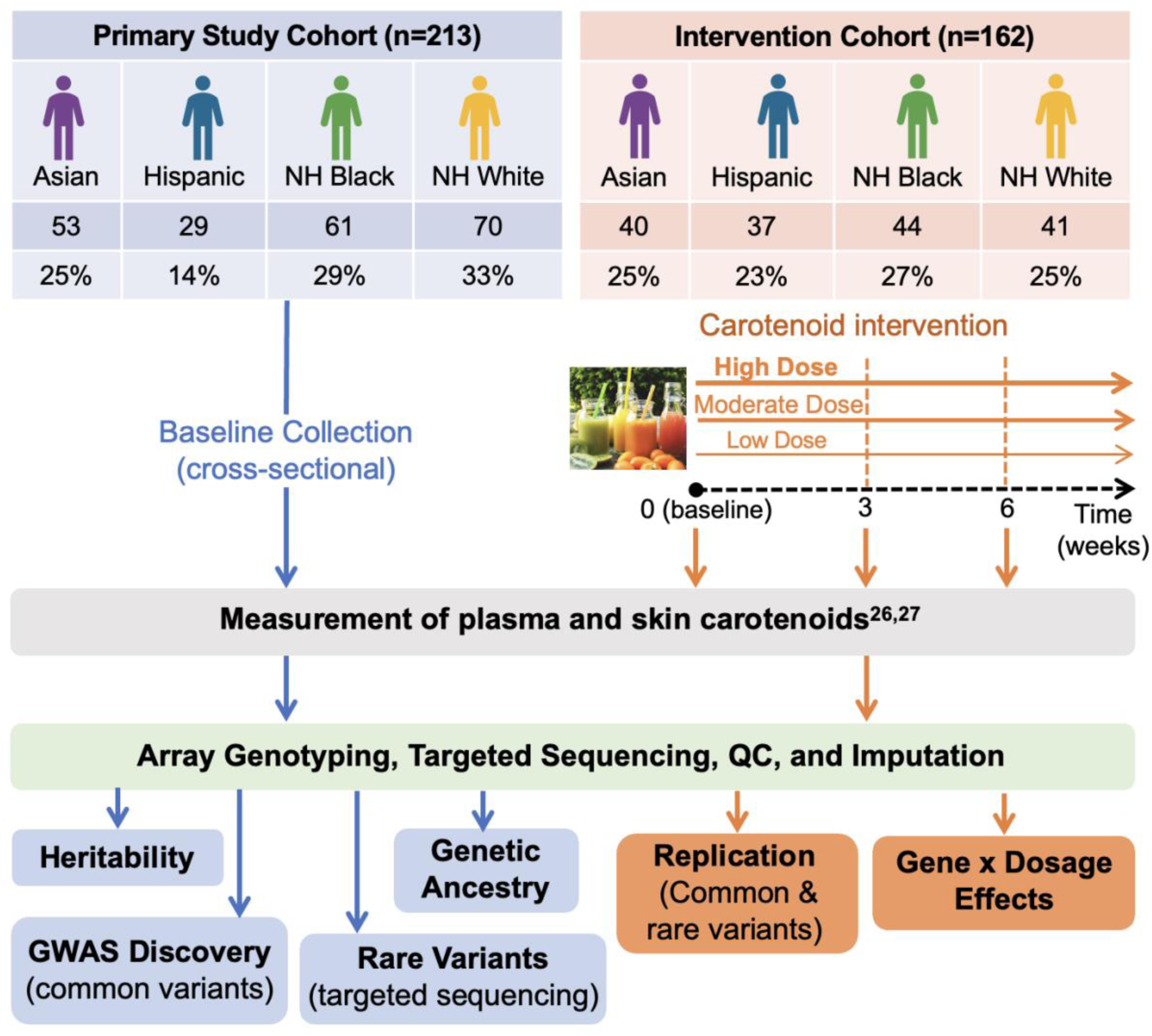
Overview of study design and analysis workflow. The study includes participants from two cohorts: the **Primary Study Cohort** (n=213) and the **Intervention Cohort** (n=162), both comprised of multiple racial/ethnic groups (Asian, Hispanic, non-Hispanic Black, and non-Hispanic White), with corresponding sample sizes and proportions detailed in the top panel. The Intervention Cohort was divided into three groups based on fruit and vegetable (FV) intake: Low Dose/Control (negligible carotenoid intake), Moderate Dose (4 mg total carotenoids/day), and High Dose (8 mg total carotenoids/day). Baseline measurements of plasma and skin carotenoids were performed for all participants. For the Intervention Cohort, additional measurements were collected at weeks 3 and 6 post-intervention. Data were collected from previous studies (see Reference 6 and 8). Array Genotyping data from the Study Cohort was used for genetic ancestry assessment and heritability estimation, stratified by race/ethnicity. Both common and rare variants identified in the Study Cohort were further analyzed for interaction effects in the Intervention Cohort.

**Figure 2.**
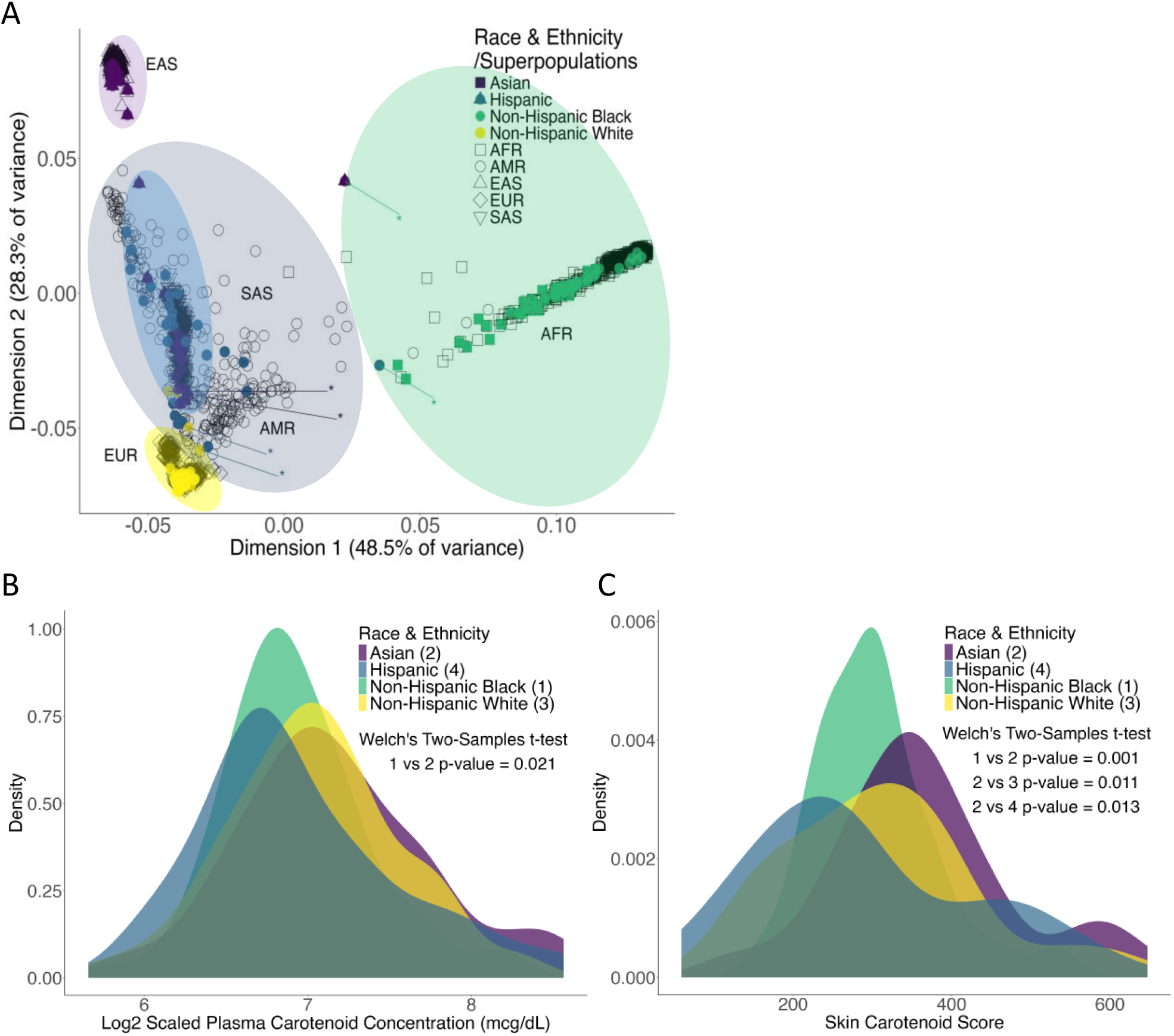
Self-reported and genetically defined race/ethnicity and total carotenoid concentrations in primary cohort. Self-identified race/ethnicity groups include Asian (n=53), Hispanic (n=29), Non-Hispanic Black (n=61), and Non-Hispanic White (n=70). **2A** - Multidimensional Scaling (MDS) of genomic data alongside ‘superpopulations’ from 1000 Genomes Project: AFR (African), AMR (Admixed American), EAS (East Asian), EUR (European), SAS (South Asian). ‘*’ indicates individuals whose self-reported ancestry differs from genomic alignment. **2B** - Plasma carotenoid concentrations (mcg/dL, log_2_-transformed); **2C** - skin carotenoid scores. Group number is indicated in parentheses, and differences were assessed via Welch’s Two-Sample *t*-tests.

### Ancestral background influences carotenoid phenotype variability

Carotenoid concentrations in plasma and skin are influenced by a variety of factors, including indistinct biological factors proxied by self-reported racial-ethnic background^27,28^. Overall, we did not observe significant variation in the plasma carotenoid concentrations and skin carotenoid levels among the four race/ethnicity groups in the Primary Study Cohort (ANOVA); however, several significant differences in total plasma carotenoid levels were observed among pairwise comparisons between the four primary cohort groups. Self-identified non-Hispanic black and Asian individuals showed the most significant differences, including in total plasma carotenoid (Welch’s Two-Sample *t*-tests P = 0.021), plasma β-carotene (P = 0.003), and plasma lutein/zeaxanthin (P = 0.037) (**Figure 2B**; **Supplementary Figure S2**). Additionally, levels of α-carotene, lycopene, cryptoxanthin, and skin carotenoids differed significantly between all four groups, with *t-*test p-values ranging from 8.67E-5 (non-Hispanic black vs. Asian for α-carotene) to 0.031(non-Hispanic black vs. non-Hispanic white for lycopene) (**Figure 2** and **Supplementary Figure S2**), while without significant differencs between the EAS and SAS (**Supplementary Figure S3**). Correlations between skin and plasma carotenoids were not different between self-reported race and ethnicity^27^; however, significant pair-wise differences in skin carotenoid levels between self-reported groups mirrored observations in plasma carotenoids, although the variance in the distributions of carotenoid species in groups was broader (**Figure 2C**).

The Primary Study Cohort included self-identified racial and ethnic groups consistent with historical US census race and ethnicity categories; to appropriately contextualize these groups for genetic studies, we aligned recruited individuals to genetic ancestry superpopulation clusters from the 1000 Genomes Phase III dataset^29^ using multi-dimensional scaling (MDS). Most individuals identifying as ‘white’ and ‘non-Hispanic black’ clustered closely with ‘European’ and ‘African’ genetic ancestry superpopulations, respectively. The reported ‘Asian’ race and ethnic group, however, separated into two distinct clusters on the first two dimensions of the MDS, with some individuals clustering with Indian/South Asian individuals (GIH) and others aligning with East Asian ancestral groups (JPT, CHB) (**Figure 2A**). Individuals self-identifying as ‘Hispanic’ clustered with mixed American and Hispanic ancestral groups (AMR). A small group of individuals displayed notable discrepancies between their self-reported race/ethnicity and genetic clustering (**Figure 2A**), including four individuals reported as ‘white’ and two reported as ‘Asian’ whose genetic ancestry aligned more closely with ‘South Asian’ or ‘Hispanic’ and ‘AFR’ super populations, respectively. (**Figure 2A**). For individuals discordant between self-reported and genetically clustered ancestry, their genetic ancestry was used in downstream analyses.

### The heritability of carotenoid concentration varies by carotenoid species and genetic ancestry

To evaluate heritability in our cohort and facilitate downstream genetic analyses, we derived a curated, quality-controlled (QC) dataset of 1,917,156 genotyped single nucleotide polymorphisms (SNPs) from 207 healthy, unrelated individuals (**Supplementary Methods**). This dataset was used to impute a total of 26,084,710 SNPs utilizing the Michigan Imputation Server^30^; with the 1000 Genomes Phase III v5 (GRCh37/hg19) reference panel serving as the primary reference for this cohort of diverse US individuals. Further filtering for SNPs with a correlation coefficient (r^2^) of 0.3 or greater, and a minor allele frequency (MAF) greater than 0.05 resulted in a final dataset of 7,467,403 SNPs. Relative and absolute heritability of total and sub-speciated plasma carotenoids, as well as skin carotenoid levels, was then calculated in GCTA^31^(**Methods**) using QCed autosomal SNPs.

In our primary cohort, the overall estimated heritability of plasma carotenoids was low (h^2^=0.08) albeit with a wide standard error (se= 0.157); perhaps unsurprising given the substantial dietary contribution to the variance in carotenoid levels. Plasma lutein/zeaxanthin had the lowest heritability estimate (h^2^=0.09, se= 0.172) among carotenoid species, also reflecting a heavy dietary influence. There was, however, notable variation in heritability estimates between carotenoid species; for instance, plasma cryptoxanthin (h^2^=0.44, se= 0.322) and *α*-carotene (h^2^= 0.35, se= 0.338) had higher heritability estimates compared to other species, which were generally <0.17 (**Figure 3A, Supplementary Table S4**). By contrast, the heritability of skin carotenoids was considerably higher than that of total plasma carotenoids (h^2^=0.08 for plasma carotenoids versus h^2^= 0.30 for skin carotenoids) and more consistent with estimates for *α*- carotene (**Figure 3A, Supplementary Table S4**).

**Figure 3.**
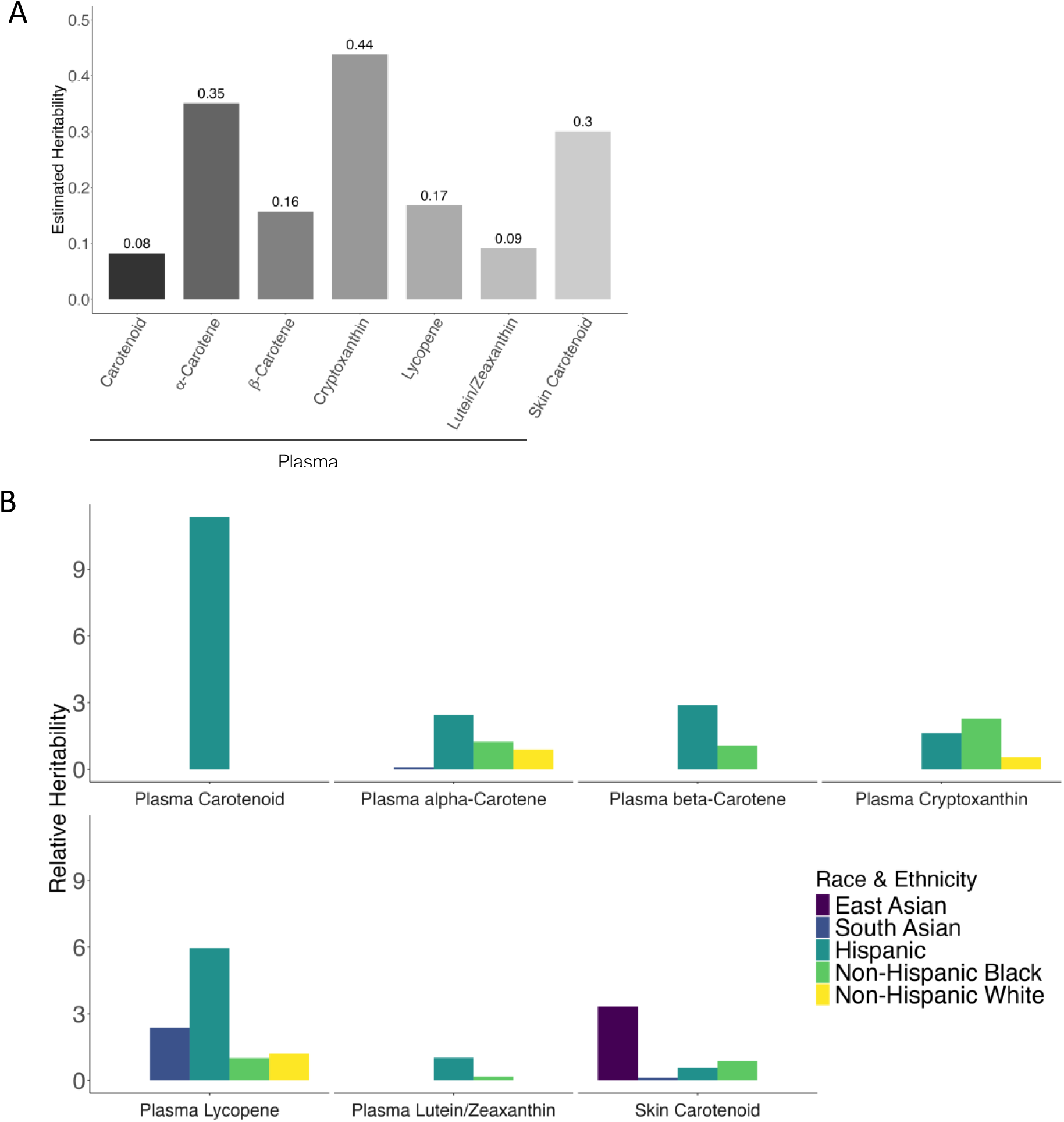
Heritability of plasma and skin carotenoids. **3A** - Heritability estimates for plasma carotenoid concentrations and skin carotenoid score across the entire Primary Study Cohort. **3B** - Relative heritability of plasma carotenoid subspecies for race/ethnicity groups. Genetic ancestry is defined based on multidimensional scaling (MDS) analysis. Relative heritability is calculated as the heritability estimate in each subgroup divided by the heritability estimate for the entire Primary Study Cohort for each carotenoid species.

To better understand the contribution of genetic ancestry to interindividual variation in carotenoid phenotypes, we also estimated heritability across each genetic ancestry group. Given the modest size of the resulting sub-samples, which resulted in large standard errors, we focused on heritability estimates relative to the entire Primary Study Cohort. The heritability of total plasma carotenoids was found to be relatively consistent across groups, except among participants genetically clustering with Hispanic individuals (**Figure 3B**), in whom it was 9x higher. A similar trend was observed for plasma α-carotene, β-carotene, and total lycopene, which all exhibited higher relative heritability among Hispanic (AMR) clustering participants (**Figure 3B**). By contrast, plasma cryptoxanthin had a relatively higher heritability (2.3x) among African American clustering individuals (AFR), while the heritability of plasma lycopene was notably higher among South Asian (SAS) clustering individuals (2.4x). The heritability of skin carotenoids was highest among East Asian (EAS) clustering individuals (3.3x) (**Figure 3B**). Generally, ancestry-specific relative heritability for skin carotenoids was higher among groups outside of the European genetic cluster (**Figure 3B**).

### Common variants at novel loci are associated with plasma, but not skin, carotenoid concentrations

Given the relatively high heritability of some of the carotenoid species, we next sought to identify genetic loci with significant effects on carotenoid concentrations across our diverse cohort. Carotenoid measurements were log_2_-transformed to provide a better approximation of a normal distribution to be used in our statistical models (**Supplementary Table S3**). For most measurements, such as plasma and food carotenoids, the log_2_-transformed values showed improved normality (e.g., for food carotenoids, W = 0.794; P = 9.691E-16 in the original data, W = 0.993 and P = 0.422 after log_2_ transformation) (**Supplementary Table S3**). However, for skin carotenoid measurements, the original values had a better fit to normality. Using the transformed plasma values, we first conducted a genome-wide association study (GWAS) of total carotenoids and carotenoid species concentrations in plasma using linear regression models as implemented in GEMMA and incorporating covariates of age, sex, BMI, log_2_-transformed carotenoid intake, and the first two MDS dimensions (**Supplementary Figure S1**).

In our analysis of plasma carotenoids, we identified six SNPs at three loci that reached genome-wide significance (P = 5E-08) for either total- or plasma carotenoid species (**Table 1**, **Figure 4**). A total of 37 SNPs at 12 loci surpassed a more permissive suggestive association threshold (P < 5E-06). The strongest association was observed between plasma α-carotene concentrations and rs3765544 (chr9:134458148:G>A) on chromosome 9q34 (P = 8.86E-10, beta = 0.750; **Table 1**, **Figures 4B & 4G**), located in the intronic 24 region of the *RAPGEF1* gene. This SNP had an effect size translating to an increase of 68% more α-carotene concentration per allele. *RAPGEF1* encodes a guanine nucleotide exchange factor involved in the activation of *Ras* family GTPases^32,33^ that plays a role in several cellular signaling pathways^34,35,36^, and it is widely expressed in tissues such as skeletal muscle and adipose tissue. Notably, the same SNP allele (G) at this locus also showed marginal association (P = 6.43E-08, beta = 0.789) with β-carotene levels (**Table 1**, **Figure 4C & 4F Supplementary Table S5**), suggesting shared activity.

**Figure 4.**
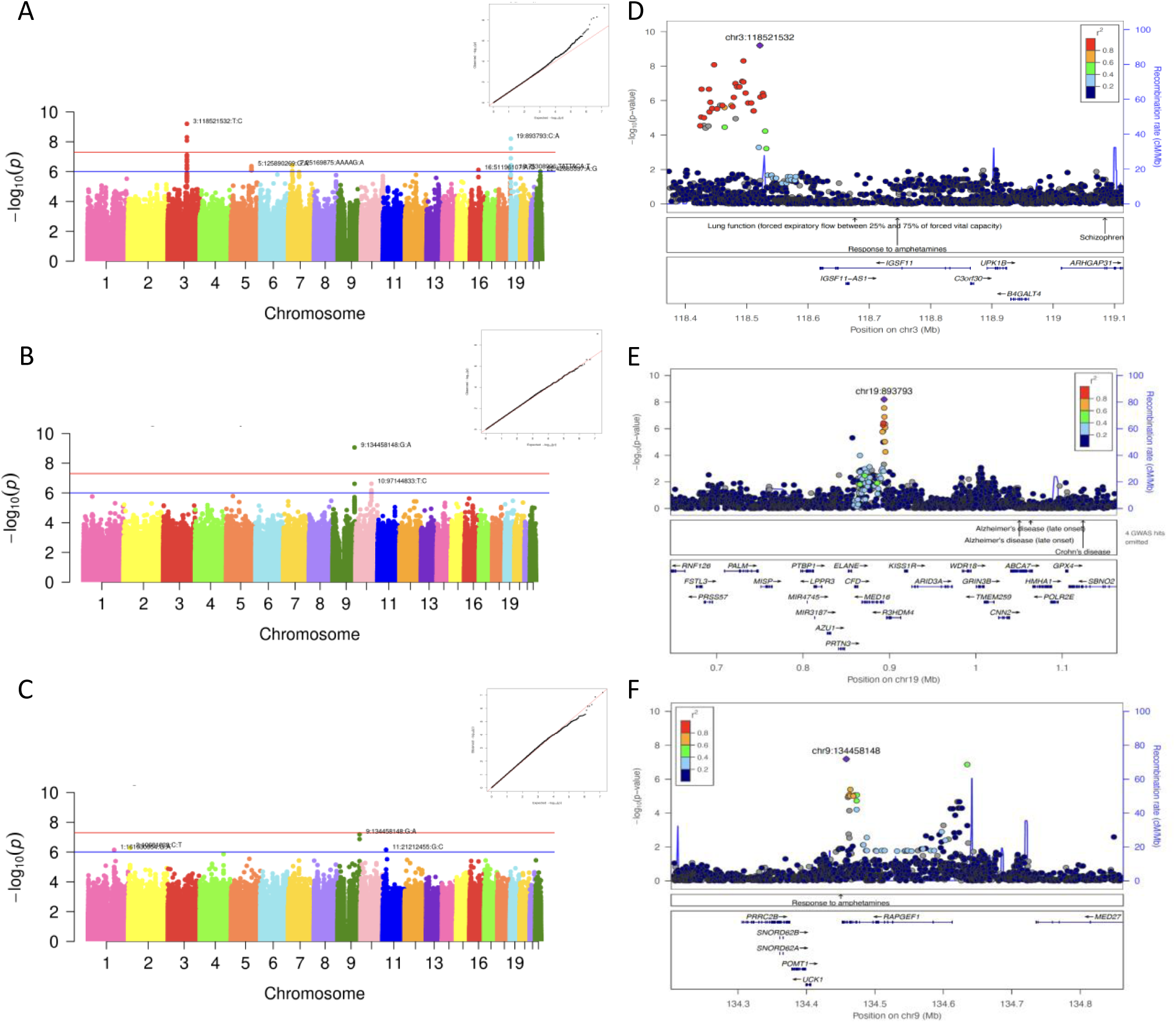
Genome-wide association analysis of plasma carotenoid and subspecies concentrations. Manhattan and Q-Q plots from GEMMA linear regression analysis of log_2_- transformed plasma cryptoxanthin (**4A**), α-carotene (**4B**), and β-carotene (**4C**). Genome-wide significance is indicated by red line (-logP = 7.3) and suggestive association by the blue line (- logP = 6). LocusZoom plots of SNP associations with carotenoid levels at significant loci: 3q13 (*IGSF11)* (**4D**), 19p13 (*R3HDM4/MED16)* (**4E**), and intragenic to *RAPGEF1* (**4F**). Recombination rates and linkage disequilibrium (r^2^) are relative to the AFR superpopulation in 1000 Genomes.

**Table 1.** SNPs significantly associated with carotenoid concentration phenotypes. SNPs in bold were directly genotyped on the Infinium™ H3Africa Consortium Array v2; the remaining SNPs were imputed. SNP ID includes chromosome: chromosome position: reference allele: alternate allele. Phenotypes are log₂-transformed carotenoid concentrations in plasma. AF-Minor Allele Frequency; Beta–logistic regression slope (effect size); P - p-value from the GEMMA analysis; **P_Met_**_a_ - p-value from the METALysis. Dir – direction of effect (positive or negative).

| Phenotype<br>(Plasma) | Nearest<br>Gene | SNPs | rsID | Primary Study Cohort |  |  | Intervention<br>Cohort |  |
| --- | --- | --- | --- | --- | --- | --- | --- | --- |
|  |  |  |  | AF | Beta | P | P<sub>Meta</sub> | Dir |
| **α-Carotene** | ***RAPGEF1*** | **9:134458148:G:A** | **rs3765544** | **0.054** | **0.75** | **8.86E-10** | **1.74E-09** | **+?** |
| **β-Carotene** | ***RAPGEF1*** | **9:134458148:G:A** | **rs3765544** | **0.054** | **0.79** | **6.43E-08** | **1.01E-07** | **+?** |
| Cryptoxanthin | *IGSF11* | 3:118446886:T:C | rs76087842 | 0.08 | 0.72 | 8.32E-09 | 1.32E-06 | -- |
| **Cryptoxanthin** | ***IGSF11*** | **3:118492749:G:A** | **rs798600** | **0.129** | **0.55** | **7.63E-08** | **3.52E-06** | **--** |
| Cryptoxanthin | *IGSF11* | 3:118494728:C:G | rs76613159 | 0.078 | 0.71 | 4.95E-09 | 1.62E-08 | -- |
| Cryptoxanthin | *IGSF11* | 3:118494740:T:C | rs1088589 | 0.137 | 0.53 | 8.21E-08 | 1.08E-04 | +– |
| Cryptoxanthin | *IGSF11* | 3:118521532:T:C | rs80316816 | 0.08 | 0.74 | 6.25E-10 | 1.34E-06 | –+ |
| Cryptoxanthin | *R3HDM4* | 19:893793:C:A | rs28468554 | 0.359 | -0.46 | 6.26E-09 | 1.76E-06 | -- |
| **Cryptoxanthin** | ***R3HDM4*** | **19:893910:A:G** | **rs28626160** | **0.495** | **-0.37** | **2.77E-08** | **1.96E-05** | **+–** |

Consistent with the high heritability seen for plasma cryptoxanthin, we observed two genome-wide significant loci associated with plasma cryptoxanthin concentrations: 1) multiple SNPs downstream of *IGSF11* on chromosome 3q13 (**Figure 4A & 4D**; top SNPs: chr3:118521532:T>C (rs80316816), P=6.25E-10, beta=0.08; chr3:118494728:C>G (rs76613159) P=4.95E-09, beta=0.08; chr3:118446886:T>C (rs76087842) P=8.32E-09, beta = 0.08) and 2) an intergenic locus on chromosome 19p13, comprising multiple SNPs (**Figure 4E**; top SNPs: chr19:893793:C>A (rs28468554), P=6.26E-09, beta=-0.462). Collectively, these two loci account for approximately 32.95% of the variance in cryptoxanthin concentration. The 3q13 locus includes (within 50kb) genome-wide significant SNPs associated with education attainment in two large studies^37,38^ and may not be regulatorily related to the closest gene (*IGSF11*). In the Genotype-Tissue Expression (GTEx) database^39^ the top SNP on 19p13 (rs28468554) is an expression quantitative trait locus (eQTL) SNP for *MED16* in multiple tissues. *MED16* encodes a protein of the same name that is a component of the mediator complex^40^, which enables thyroid hormone and vitamin D3 receptor binding^41,42^.

We used the same GWAS model to evaluate SNPs associated with skin carotenoids, but for this analysis, we also included measurements of melanin and hemoglobin - both of which are thought to influence skin carotenoid measurements^27^ - as covariates. No variants surpassed either the genome-wide or suggestive association threshold. We considered that by including skin tone measurements and accounting for ancestry (as modeled in GEMMA), we may have overcorrected for the ancestry effect; that is, if melanin and hemoglobin are collinear with some genetic ancestries, also incorporating components reflecting genetic ancestry could be redundant. Therefore, we reran the association using only clinical covariates (i.e. without MDS coordinates or race/ethnicity) (**Supplementary Figure S4A**). This yielded 107 SNPs surpassing our genome-wide significance threshold (**Supplementary Figure S4B,** genomic inflation factor = 2.06); this suggested a strong effect of ancestry (population stratification) and was reflected in disparities in minor allele frequencies at ‘associated’ loci between different genetic ancestry groups (**Supplementary Figure S4D**). This disparity in association with and without ancestry adjustments was not observed in the plasma carotenoid association analyses (**Supplementary Figure S4C**).

### Replication and gene-by-dosage effects of carotenoid candidate SNPs

To replicate our primary cohort findings, we conducted genome-wide genotyping in a secondary cohort derived from a dietary carotenoid intervention study with a similar study design^26^ (**Methods**). This secondary cohort was smaller in size (n=162) and had a larger proportion of Hispanic clustering (AMR, 23% vs. 14%), and a smaller proportion of European clustering (EUR, 27% vs. 33%), individuals relative to our initial cohort (**Figure 1**). The distribution of sex in the second cohort (male: n=79, 49%; female: n=83, 51%) is nearly equal, contrasting with the predominantly female first cohort (male: n=62, 29%; female: n=151, 71%). while age (median age = 29 years) was not different between the two cohorts (**Supplementary Table S1 & S2**). Secondary cohort samples were genotyped on the same platform and underwent identical quality control procedures as the primary cohort. We used baseline (pre-intervention) plasma carotenoid and species measures as the outcome phenotype, applying the same GWAS covariates and linear regression models in GEMMA with the 110 non-overlapping individuals (i.e. only unrelated individuals who did not participate in the primary cohort were included in analyses).

Meta-analysis between the two cohorts was then conducted in METAL^43^ for the 37 suggestive threshold SNPs (representing 12 loci). We found nominal evidence for replication of the same carotenoid species (cryptoxanthin) at the same two loci noted in our primary analysis (*IGSF11* at 3q13 and *RNF111/MED16* at 19p13). Meta-analysis of rs1088589 at *IGSF11* surpassed genome-wide significance, with a similar direction and magnitude of effect in both cohorts (p = 9.69E-08, beta = 0.532) (**Table 1**). Additionally, several other SNPs in this region (8 out of 12) surpassed the suggestive meta-analysis threshold (P < 1E-06) and exhibited the same direction of effect. Two imputed SNPs, including our top SNP in *RAPGEF1*, were not observed (imputed) in the secondary cohort and thus could not be replicated.

The interventional study design in our secondary cohort involved randomizing participants to receive low, moderate, or high doses of dietary carotenoids via a daily, carotenoid-enriched, fruit and vegetable juice, with skin and plasma carotenoid concentrations measured at baseline (time 0) as well as 3- and 6-weeks post-intervention. This study design allowed us to investigate whether SNPs at candidate loci identified in our primary cohort might also influence the accumulation of carotenoids over time. We applied linear regression models (**Methods**), incorporating the same covariates as in our initial study, along with additional terms for intervention time points and a gene-by-dosage interaction effect (**Methods**). Of the 37 candidate SNPs identified in the discovery cohort and evaluated in the Intervention Cohort (**Supplementary Table S5 and S6**), 32 (86.5%) had significant F-statistic p-values (<0.05), indicating a statistically significant relationship with the outcome variable (plasma carotenoid concentration). Additionally, six (16.2%) SNPs – upstream of *IGSF11* (n=5) associated with cryptoxanthin exhibited gene-by-dosage effects, where the effect of genotype on plasma carotenoid concentrations differed by intervention dosage (**Figure 5A-F, Supplementary Figure S5**).

**Figure 5.**
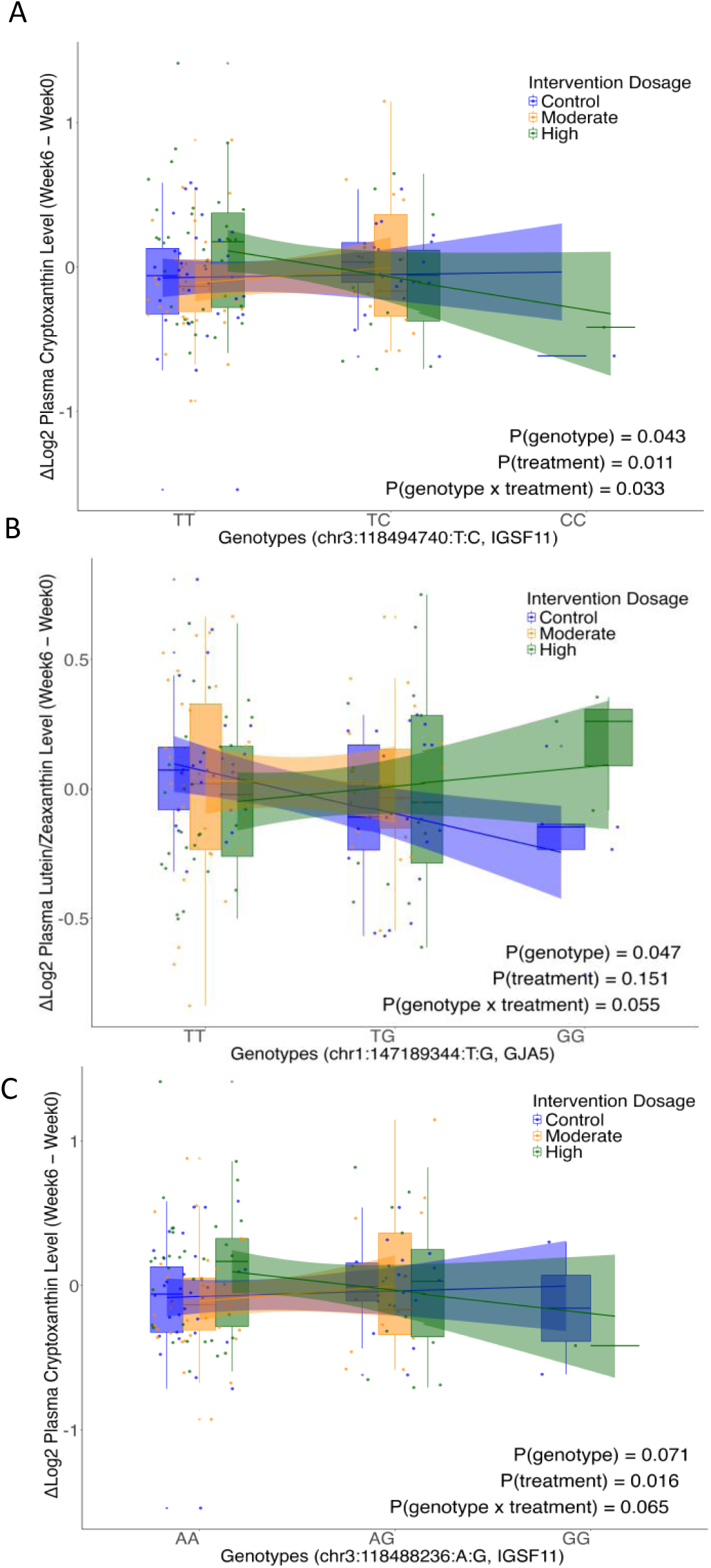
Gene-by-dosage plots. Genotypes are shown along the x-axis, with intervention dosages color-coded: blue - Control, orange - Moderate, and green - High. Box plots represent plasma carotenoid distributions within each genotype and dosage. Smoothed regression lines show genotype effects within treatments. P-values indicate the significance of genotype (P(genotype)), treatment (P(treatment)), and genotype-by-treatment interaction (P(genotype × treatment)). Treatment p-values < 0.1 indicate significant dosage effects.

### Target sequencing captures association between PKD1L2 and **β**-carotene concentrations

Previous genome-wide studies have demonstrated an association between β-carotene and common variants upstream of β-carotene oxygenase 1 (*BCO1*)^12^. However, our primary GWAS dataset lacked strong evidence for this. Whilst population differences (Eurocentric vs diverse cohort) and sample size undoubtedly underlie part of this observation, we also noted low coverage (few polymorphic SNPs) in this region on the genotyping array used; this is a known limitation of array-based studies in genetically diverse populations^44,45^. To address this, we performed targeted sequencing of a subset of candidate genes (**Supplementary Table S7**) identified from the literature^4,46,47^ and not adequately covered by the array. This sequencing provided consistent median coverage across targeted loci (chr16:81101012-81220480), with *PKD1L2*, upstream of *BCO1*, exhibiting a higher number of variants than other genes (**Supplementary Figure S7**).

*PKD1L2* single nucleotide variants (SNVs) were the only SNVs from our targeted sequencing cohort that were consistently associated with plasma or skin carotenoid levels in our linear regression models, specifically with β-carotene concentrations. This association remained significant when comparing the top third versus the bottom third of β-carotene concentrations (**Supplementary Figure S7**). *PKD1L2* variants also showed the strongest associations in the replication cohort, with a similar direction (positive association) and effect sizes across the two cohorts, though the specific variants differed (**Supplementary Figure S7**). The *PKD1L2* locus is found upstream of *BCO1*, in a region consistently associated with β-carotene concentrations. Although our targeted capture did not directly sequence previously reported SNPs upstream of *BCO1*^11^, linkage disequilibrium (LD) patterns suggest that *PKD1L2* variants are likely to be in the same LD block. As the *BCO1*-*PKD1L2* association was initially observed among the European genetic ancestry group, we evaluated genotypes and β-carotene levels across our four genetic ancestry groups. The top two SNPs (rs4148211 and rs7194871) were analyzed for associations with β-carotene concentrations across four ancestry groups. A nominally significant association was observed for rs4148211 in the Asian group (P=0.012, n=51), while no significant associations were found for rs7194871 (p>0.5).

### Rare, protein-damaging variants are observed in individuals with outlier carotenoid concentrations

Rare, protein-damaging variants in coding regions of genes can have large effects on physiologic traits^48^. We, therefore, looked for rare, putatively protein-damaging variants (**Supplementary Methods**) among individuals with extreme values (>2SD or <2SD) for either plasma or skin carotenoids across Primary Study Cohort (**Supplementary Table S8**).

In the Intervention Cohort, we identified an individual carrying a rare, predicted-damaging, missense coding variant (**Supplementary Table S9**) (rs142824860, NC_000016.9:g.81272554A>G; p.Glu14Gly; gnomAD MAF = 9.7E-05; CADD score 25.0) (**Supplementary Methods**) in the *BCO1* gene who also had the lowest plasma β-carotene concentrations in the cohort. For skin carotenoid concentrations, three notable outliers were observed: two of these individuals clustered with the 1000 Genomes EAS population, and both individuals carried a missense coding variant in *CETP* (rs2303790, NM_000078.3:c.1376A>G; p.Asp459Gly; gnomAD MAF = 0.002) that is seen in 3% of East Asian (EAS) clustering individuals but <1% in all other populations. The third outlier, clustering with other European (EUR)-identifying individuals, possessed an ultra-rare (MAF=9.9E-06) variant in *APOA1* (rs756535387; p.Arg201Ser; CADD score 24.0), predicted to be deleterious with an AlphaMissense score of 0.701 (likely pathogenic) (**Supplementary Methods**). All three outliers also had elevated plasma cryptoxanthin concentrations, which were strongly correlated with skin carotenoid levels (r = 0.57; **Figure 6**).

**Figure 6.**
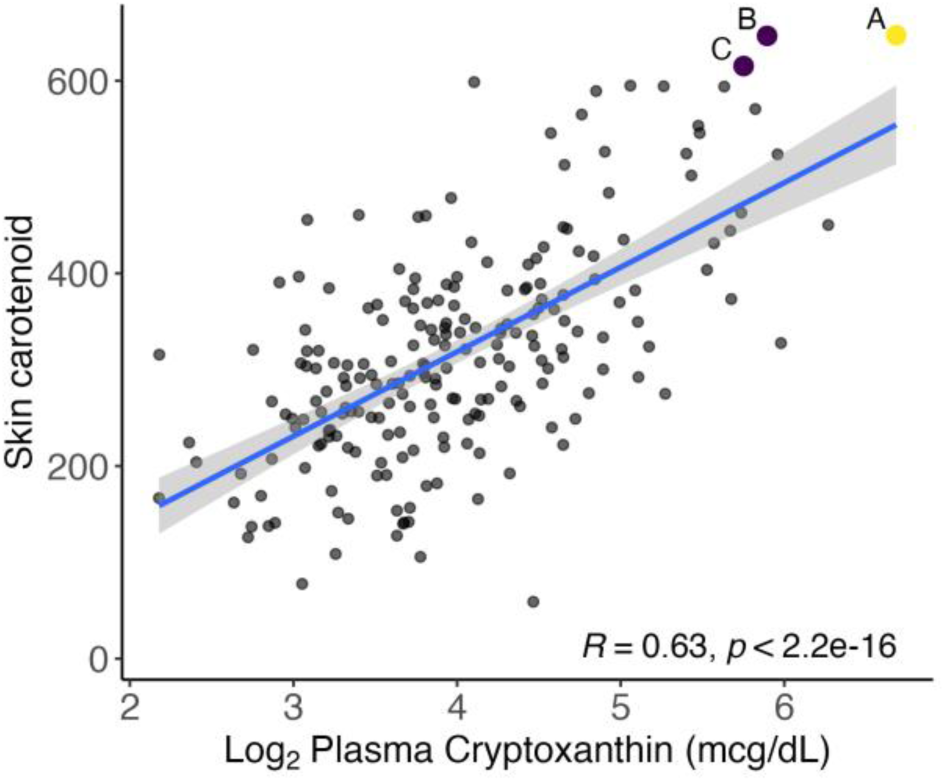
Correlation between skin carotenoid levels and plasma cryptoxanthin. Scatterplot shows skin carotenoid levels (y-axis) and log_2_-transformed plasma cryptoxanthin concentrations (x-axis). Each gray dot represents a participant. The blue line represents the linear regression fit, with a significant positive correlation (R = 0.63, p < 2.2 x 10^-16^). Outliers (purple; B and C), who self-identified and clustered by genomic information as Asian, carry the rare East Asian-specific *CETP* variant (rs2303790). Outlier (yellow; A), self-identified and clustered by genomic information as Non-Hispanic White, has a rare *APOA1* variant (rs756535387) predicted to be deleterious. Details of deleterious variants are given in **Supplementary Table S9**.

## DISCUSSION

Through GWAS and targeted sequencing analyses in two ancestrally diverse cohorts, we provide comprehensive estimates of heritability and the genetic architecture underlying plasma and skin carotenoid concentrations. We find that heritability varies across plasma carotenoid species and is influenced by genetic ancestry, with estimates being generally lower for plasma carotenoids compared to skin carotenoids. Notably, we identify and replicate an association between genetic variation at chromosomes 3q13 (upstream of *IGSF11*) and 19p13 (*MED16*) and plasma cryptoxanthin concentrations and find evidence of gene-by-dosage effects at the 3q13 locus. We confirm the association between β-carotene at the *PKD1L2*-*BCO1* locus and identify putatively protein-damaging rare variants with large effects on skin and plasma carotene concentrations.

We observed substantial heritability for plasma carotenoid species α-carotene and cryptoxanthin, as well as skin carotenoids, with estimates comparable to those observed for serum lipids such as cholesterol (ranging from 0.30 to 0.70)^49,50^; this is consistent with the shared biochemistry and metabolism of serum lipids and carotenoids. We also noted significant variation in heritability estimates across different genetic ancestry groups, with individuals clustering with admixed Amerindians (AMR; self-identified ‘Hispanic’) exhibiting high heritability for nearly all plasma carotenoid sub-species, including replication of the high heritability of α-carotene in previous studies among Mexican Americans (h^2^ = 0.85 in this study; 0.81 in the previous study)^8^. Given that heritability represents the genetic contribution to total variance, these results likely reflect the balance between genetic variation and differences in typical dietary and environmental factors across groups. That some carotenoid species have higher heritability estimates suggests that the metabolic events preceding their plasma accumulation, such as intestinal absorption, are more sensitive to genetic variation. The public health implication thereof is that dietary recommendations (or interventions) for certain carotenoids (e.g. cryptoxanthin) aimed at achieving a specific plasma concentration range may be more challenging to develop than for those carotenoid species with lower heritability. Larger sample sizes will be important in further elucidating these differences and their public health implications.

The distinct contributions of diet and genetics were further illustrated in our analyses of skin carotenoid heritability. Non-invasive measurements of the tissue accumulation of carotenoids in the skin present unique considerations. Principally, the efficiency of detecting colorimetric changes due to carotenoid accumulation in the skin depends partially on skin reflectivity, which may be influenced by skin melanin content and hemoglobin levels. Previously, we demonstrated that, after adjusting for melanin, hemoglobin, and dietary intake, a robust correlation remains between skin and plasma total carotenoid concentrations^27^, though we subsequently did not find skin melanin and hemoglobin to be significant modulators of skin carotenoid responses to changes in dietary carotenoid intake^10^. Consequently, the apparent heterogeneity in heritability estimates across ancestry groups could be a combination of variability in confounding skin parameters and ancestral variability in factors influencing tissue accumulation. The latter contention is worthy of consideration given the importance of carotenoids to vitamin A metabolism, as carotenoids serve as precursors to retinoids, which are critical for maintaining skin health^46^. This highlights the potential influence of ancestral variability on vitamin A-related physiological processes and the tissue accumulation of carotenoids. Despite this, our understanding of the factors influencing tissue carotenoid accumulation remains limited and warrants further investigation.

The relatively higher heritability for plasma cryptoxanthin was reflected in the number of genome-wide- and suggestive associations compared to concentrations of other carotenoid species, especially at the 3q13 and 19p13 loci. Few if any previous genetic studies have considered cryptoxanthin concentrations, and to the best of our knowledge, a role for genetic variation at these two loci has not previously been described. At 3q13, the closest gene, *IGSF11*, encodes a member of an immunoglobulin superfamily^51^ whose primary function is as a cell adhesion molecule that stimulates cell growth^52,53^; however, in addition to neurocognition, genetic variation at 3q13 has been implicated in gut microbiota diversity^54,55^ and body mass index^56–58^, suggesting that the full functional regulatory impact of this locus may not yet be well understood. The 3q13 locus also provided five of the six SNPs (all in strong linkage disequilibrium (LD) with each other) with evidence for gene-by-dosage effects, with the mutant (non-reference) minor allele being negatively correlated with cryptoxanthin concentrations at high FV doses but being positively correlated or neutral at low or intermediate FV doses. We documented the gene-by-dosage effect for dietary carotenoid intervention and further highlighted the complexity of making personalized nutrition recommendations for specific carotenoid species. Cryptoxanthin, while a relatively small component of total plasma carotenoids, serves as a highly bioavailable vitamin A precursor, and confers reduced inflammation, improves immune function, and antioxidant activity^59–61^. A recent longitudinal population study found a strong positive association between maternal cryptoxanthin concentrations at delivery and offspring cognitive development at age two^62^.

Targeted sequencing further enhanced our ability to capture the full spectrum of genomic variation, particularly given the diverse ancestries included in our study. This approach facilitated the interrogation of loci that are not well captured or imputed in diverse cohorts using genotyping microarrays, and the identification of rarer and novel variants that would not be detectable from fixed-content arrays. This was most evident at the *PKD1L2*-*BCO1* locus; these two genes are arranged in reverse tandem (head to tail) within an ∼18 kb stretch on chromosome 16q. Common variants in this region – ranging from the 5’upstream of *BCO1* to *CETP* have been consistently associated with β-carotene metabolism^11,12,63^; however, narrowing down putatively causal variants in this region has been elusive. Our findings underscore this uncertainty –the top associated variants were in the fifth exon of *PKD1L2*, but the top associated variant was much further downstream of *PKD1L2* in our Intervention Cohort (**Supplementary Figure S7**). The challenge of replicating individual variants at this locus across studies likely stems from differences in population ancestry, environmental factors (such as adequately accounting for dietary carotenoid intake), and study design, all of which may distort the association of variants, particularly if multiple associated alleles each have small effect sizes. Regardless, the gene-level association is sufficiently consistent that the *PKD1L2* may harbor multiple common variants, each contributing modestly to β-carotene levels, collectively exerting a significant effect. In line with this, we identified a rare, damaging variant (rs142824860) in the *BCO1* gene in our Intervention Cohort in an individual with the lowest β-carotene levels, further highlighting the potential for multiple variants with varying effects to contribute to population levels of β- carotene^10^.

Our results also suggest a strong putative overlap between lipid metabolism and physiological carotenoid regulation. Six genes (*ALDH7A1*, *ATF6*, *MED16*, *SALL1*, *SORBS1*, *SORBS2*) near our plasma carotenoid suggestive candidate loci, as well as both genes (*CETP* and *APOA1*) harboring high-impact rare variants in individuals with outlier carotenoid concentrations, are either known or suspected modulators of lipid metabolism^64–69^. Among these, the *ATF6* locus on chromosome 1had the strongest statistical association, with the rs11579627 SNP (chr1:161930954:G>A) nearing genome-wide significance (P = 8.23E-08, beta = 0.32), **Supplementary Table S5**). Notably, ATF*6*, a key transcription factor in the endoplasmic reticulum (ER) stress response and the unfolded protein response (UPR) pathway^70^, is implicated in lipid biosynthetics^65^. These associations highlight allelic variation in genes predominantly involved in cell signaling and lipid metabolism.

The missense coding variant in *CETP* has been previously linked to exudative age-related macular degeneration^71,72^ (a condition related to carotenoid nutrition) and elevated high-density lipoprotein cholesterol levels (a determinant of plasma carotenoid concentrations)^73^. Whilst the overlap between lipid and carotenoid loci is not entirely surprising, given similarities in the biochemistry of both, it does suggest that identifying genetic contributors to carotenoid concentrations in larger studies could benefit from overlapping a comprehensive compendium of lipid metabolism genes and that genetic studies of lipid variation, particularly in diverse populations, would benefit from including carotenoid assessments and using colocalization to identify strong biological candidates.

The modest sample sizes and relatively balanced distribution of ancestry groups in our study means that our study was necessarily aimed at identifying loci with large trans-ancestry effects that are likely to be relevant across ancestries. Conducting our analysis in a diverse multi-ancestry cohort, however, still provided unique insights that would not have been evident using a more ancestrally homogenous study group, particularly as it pertains to the allelic spectrum underlying carotenoid variability. For instance, most of the carotenoid heritability estimates were relatively higher among non-European populations, and all of the suggestive and genome-wide significant candidate variants observed had higher minor allele frequencies in non-European populations; this was particularly true for skin carotenoid concentrations. The inclusion of diverse genetic ancestry in our rare variant studies further emphasized the utility of ancestrally diverse cohorts – the coding missense variant in *CETP* (rs2303790) associated with very high skin and cryptoxanthin levels is predominantly common among individuals with East Asian ancestry.

Despite the insights gained from our analysis, there are limitations to our study. Our sample size is small in comparison to modern GWAS; whilst this undoubtedly limited our power to detect variants/loci with smaller effect sizes, our sample size is comparable to that used to discover major effect loci for more commonly measured physiologic proteins (e.g. fetal hemoglobin levels and cholesterol), and replicating our results in a second independent cohort mean that the reported associations are unlikely to be false positives. Despite the well-documented health effects of carotenoids, carotenoid concentrations are not routinely measured clinically or included in large-scale biobanks and databases; as a result, these resources are not available to further replicate our findings. Additionally, although supplemented by imputation and, for some loci, targeted sequencing, our reliance on genotyping arrays, particularly in a cohort with diverse ancestries, may have missed rare or novel variants that could either be independently associated with carotenoid concentrations or augment findings at suggestive loci. Potential differences in population structure (e.g. linkage disequilibrium) and/or nutritional factors between the discovery and replication cohorts may have contributed to a lack of replication of some discovery associations, especially if multiple associated alleles each have small effect sizes.

Going forward, there are several lessons for future carotenoid and nutrigenetic research. Principally, from a genetic standpoint, larger and more diverse cohort studies of carotenoids are necessary to replicate our findings and enhance the robustness and generalizability of the identified associations. Methods of incorporating local genetic ancestry^74^ and deconvoluting ancestry effects in trans-ancestry GWAS continue to improve and are likely to be particularly important for exploring and refining carotenoid associations, given the variability noted across ancestry groups. Additionally, incorporating detailed phenotyping of potential covariates, controlled interventions, and measurements of lipids and related physiological compounds is likely to be fruitful in understanding the complex underlying physiology. The rare variant candidates identified here provide strong starting points for *in vitro* and *in vivo* functional validation studies, particularly at the *CETP* (rs2303790) and *BCO1* genes. Finally, large population-based studies would provide the necessary data to consider developing personalized risk profiles that incorporate carotenoid (and related compound) measurements, genetic factors, and independent demographic and dietary interactions. Such studies have the potential to provide the level of detail needed to tailor public health recommendations for FV intake and interventions across different population groups.

The comprehensive, agnostic view of the genetics of carotenoid metabolism presented here provides a robust starting point for future studies of this important class of natural dietary compounds and underscores the necessity of including diverse ancestry groups and deep phenotyping in precision nutrigenetic research going forward.

## METHODS

### Study design and carotenoid species measurement

We utilized the samples and phenotypic data collected from individuals who participated in two previous studies^26,27^. In the first study, participants were healthy adults aged 18-65 years, recruited from two sites in North Carolina and Minnesota. They self-identified as Non-Hispanic Black or African American (hereafter referred to as Non-Hispanic Black), Asian, Non-Hispanic White, or Hispanic. The demographics of the Primary Study Cohort are detailed in **Supplementary Table S1**. Skin carotenoids were measured using pressure-mediated reflection spectroscopy (Veggie Meter, Longevity Link, Utah), which returns and aggregates skin carotenoid score measurements that correspond with multiple skin carotenoids^75,76^. Total plasma carotenoid concentrations were determined via an HPLC-photodiode array^26^. The resulting dataset comprised SNPs from a cohort with the following self-identified racial and ethnic distribution: non-Hispanic black (61, 29%), Asian (53, 25%), non-Hispanic white (70, 33%), or Hispanic (29, 14%). Participants were predominantly female (N=151 (71%)), with fewer males (N=62 (29%)), and a median age of 30 years. The second cohort was also drawn from a previous intervention study^26^, recruited from three sites in North Carolina, Minnesota, and Texas. The racial and ethnic distribution of the Intervention Cohort slightly differed from the primary cohort, consisting of non-Hispanic black (41, 25%), Asian (40, 25%), non-Hispanic White (44, 27%), and Hispanic (37, 23%) participants, with a near-equal sex distribution (49% male and 51% female) (**Supplementary Table S2)**. Plasma and skin carotenoid concentrations were collected at three time points (baseline/week 0, week 3, and week 6) using the same methods. Participants were randomized to receive negligible, medium-dose (4 mg total carotenoids/day), or high-dose (8 mg total carotenoids/day) of dietary carotenoids for the intervention. Daily intervention adherence was recorded by participants and non-intervention carotenoid intake was assessed with repeated 24- hour dietary recalls prior to each visit.

### Multidimensional Scaling (MDS)

Next, we combined our dataset with the 1000 Genomes phase III data to estimate the genetic ancestry of the cohort. The 1000 Genomes phase III data were acquired from https://ftp.1000genomes.ebi.ac.uk/vol1/ftp/release/20130502/ following the instructions at https://www.cog-genomics.org/plink/2.0/resources#1kg_phase3. After downloading, the data was converted to PLINK binary format, removing ambiguous SNPs (i.e. A>T/T>A and C>G/G>C SNPs that are indistinguishable at the strand level), non-AT, and non-GC SNPs. The data was pruned to remove SNPs with R^2^>0.1 in windows of 50 SNPs advancing 10 SNPs at a time across the chromosome (--indep-pairwise 50 10 0.1), after which SNP nucleotide mismatches were corrected and merged with our cohort data using similar QC filters. Subsequently, Multidimensional Scaling (MDS) was performed using the --cluster and --mds-plot options in PLINK, generating eigenvalues and eigenvectors that encapsulate the MDS dimensions and their respective scores for each individual. R version 4.3 was then used for plotting the MDS and scree plot.

### Genome-wide heritability analysis

Heritability estimates were calculated using Genome-wide Complex Trait Analysis (GCTA)^31^ version 1.94.1. The input data for the analysis comprised quality-controlled, genotyped, genome-wide SNPs with minor allele frequency (MAF) thresholds of 0.01(--autosome --maf 0.01). Total plasma carotenoids, plasma species, and skin carotenoid measurements for each individual included in the filtered genotyping dataset served as the phenotypic data for the heritability assessment. Genetic ancestry was used for downstream genomic analyses. Relative heritability is calculated as the heritability estimate in each subgroup divided by the heritability estimate in the entire cohort.

### Genome-Wide Association Studies (GWAS)

Genome-wide association studies (GWAS) were conducted to explore the relationship between imputed genotyping variants and carotenoid species levels, employing GEMMA^77^ version 0.98.5. As an orthogonal assessment, we also conducted association using PLINK 1.9; all primary results reported relate to tests done in GEMMA. For analyses of plasma carotenoid and species covariates of age, sex, BMI, race/ethnicity (self-reported or MDS-defined), and food carotenoid were used. Additionally, for the skin carotenoids association study, the melanin index and hemoglobin index were included, consistent with previous findings^28^. Before association testing, the normality of raw data, log_2_-transformed phenotypes, and covariates was assessed using the Shapiro-Wilk Test (**Supplementary Table S3**). Measurements approximating a Gaussian distribution were included in the association analysis. Log_2_- transformed values generally improved normality for plasma and food carotenoids, while original values better fit skin carotenoid data. GWAS was then conducted on both the study and intervention cohorts.

### Replication and transferability of significant SNPs

Suggestively associated variants (P = 5E-04) with plasma carotenoid species and skin carotenoids, identified from the GWAS in the Primary Study Cohort, were further evaluated in the Intervention Cohort. We conducted an association analysis on individuals who participated exclusively in the Intervention Cohort (n=110) using GEMMA, applying models similar to those used in the Primary Study Cohort. Baseline carotenoid measurements served as the phenotypic data. METALysis^43^ was used with significantly associated SNPs in this Intervention Cohort. For the second replication analysis, changes in plasma carotenoid species and skin carotenoid levels from baseline (week 0) to the intervention endpoint (week 6) were used as phenotypes, following the approach of a previous study^27^. Covariates including age, sex, BMI, baseline carotenoid concentrations, treatment assignment (0, 1, 2), study sites, and the first and second dimensions of MDS together with SNPs and SNPs * treatment assignment (0, 1, 2) were fitted into a linear regression model. For skin carotenoids, we also incorporated the melanin and hemoglobin index in the linear regression model.

### Linear regression model

- *Y* = Change in plasma carotenoid species or skin carotenoid levels
- *β_0_* = Intercept
- *β_1_, β_2_, β_3_,…, β_n_* = Coefficients for each covariate
- *X_1_*= Age
- *X_2_*= Sex (coded as a binary variable)
- *X_3_*= BMI
- *X_4_* = Baseline carotenoid levels
- *X_5_* = Treatment assignment (0, 1, 2)
- *X_6_* = Study site (coded as necessary)
- *X_7_* = First MDS coordinate
- *X_8_*= Second MDS coordinate
- *X_9_* = Melanin index (for skin carotenoids)
- *X_10_* = Hemoglobin index (for skin carotenoids)
- *SNP_i_* = Genotypic data for the i-th SNP (coded as necessary)
- *SNP_i_* × *Treatment* = Interaction term between SNP and treatment assignment

The linear regression model can be expressed as:

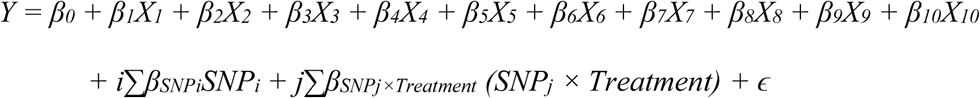

Where:

- ɛ = Error term representing the variability not explained by the model.

### Targeted sequencing and bioinformatics analysis

Thirty-five (35) genes (**Supplementary Table S7**) identified as important for carotenoid metabolism were sequenced using the Ampliseq Custom Panel from Illumina. FASTQ files were aligned to the GRCH38/hg38 reference genome using BWA^78^ MEM with the following parameters: -M -O 30 -E 4 -T 20 -v 3. Data from multiple lanes were then merged and indexed using SAMtools^79^ v1.5. Per-base coverage was computed with bedtools^80^ coverage (**Supplementary Table S7, Supplementary Figure S6**). Subsequently, the Genome Analysis Toolkit (GATK)^81–83^ v4 was utilized for variants calling following the steps of *MarkDuplicatesSpark*, *BaseRecalibrator*, *ApplyBQSR*, *HaplotypeCaller*, *GenomicsDBImport* and *GenotypeGVCFs*. SNP annotation was performed using ANNOVAR^84^ (version 2020-06-08).

### Targeted sequencing association study

The Variants Association Tool^85^ and VCFTools^86^ (v0.1.15) were employed to filter the variants within the Ampliseq target regions. Quality control filters were applied to exclude variants with a minor allele frequency (MAF) below 0.01 and a call rate below 95%. Variants significantly deviating from Hardy-Weinberg equilibrium (p < 0.001) were also excluded. We focused on non-synonymous variants by filtering out synonymous substitutions, thereby retaining only those variants with potential impacts on protein function for downstream analysis.

### Gene-by-dosage effect analysis

To evaluate the interaction between genetic variants and intervention dosage on plasma carotenoid levels, we performed a Gene-by-dosage analysis using the linear regression model above. For each locus, genotype-by-treatment interactions were assessed by fitting a model where the log_2_-transformed plasma carotenoid levels at the intervention endpoint (week 6) were the dependent variable. The independent variables included the genotype of the specific locus, intervention dosage group (control, moderate, or high), baseline plasma carotenoid levels, and a range of covariates: age, sex, BMI, study site, and race. We also included the interaction term between genotype and treatment dosage. This model was applied across loci, allowing for locus-specific examination of the gene-by-dosage effect. The significance of the gene-by-dosage effect was determined by the p-value, with a significance threshold of p < 0.1. Results were visualized using boxplots with fitted regression lines for each dosage group, illustrating the differential effects of intervention dosage across genotypes.

## Supporting information

Carotenoid Genetics Supplementary Information

Carotenoid Genetics Supplementary Tables

## Data Availability

All data produced in the present study are available upon reasonable request to the authors.

https://h3africa.org/index.php/2019/12/12/h3africa-chip-faq/

https://www.illumina.com/content/infinium-h3africa-consortium-array-data-sheet)

https://www.internationalgenome.org/

## DATA AVAILABILITY

H3Africa Array - https://h3africa.org/index.php/2019/12/12/h3africa-chip-faq/

H3Africa Array Data Sheet - https://www.illumina.com/downloads/infinium-h3africa-consortium-array-data-sheet-370-2020-001.html

1000 Genomes Project Resource - https://www.internationalgenome.org/

The datasets generated during and/or analyzed during the current study are available from the corresponding author upon reasonable request.

## ACKNOWLEDGEMENTS

The authors would like to thank all the persons who participated in data acquisition and sharing during the two cohorts’ establishment. This research was supported by the NIH NHLBI (SJP, MNL, NEM, QW: 1R01HL142544-01A1), by funding from NIH NHGRI (HG-200412 to N.A.H.), and the USDA/ARS (cooperative agreement 3092-51000-059-002S to NEM). The work and views expressed do not reflect the views of the NIH or the USDA. This work utilized the computational resources of the NIH HPC Biowulf cluster (http://hpc.nih.gov).

## AUTHOR CONTRIBUTIONS

N.E.M. and N.A.H. designed the study. M.N.L., N.E.M., and S.B.J.P. led the teams that recruited participants, obtained informed consent, and collected samples. S.M. performed the rare variant association analysis. Y.H. conducted all other data analyses, including ancestry assessment, heritability estimation, GWAS, statistical genetics, and bioinformatics processing of the target sequencing data. Q.W. provided support for statistical analysis. Y.H. wrote the initial first draft, with intellectual content added by S.M., N.A.H., and N.E.M. All authors reviewed and approved the final manuscript.

## SUPPLEMENTARY INFORMATION

### Supplementary Tables

Supplementary Table S1. Demographics of the Primary Study Cohort.

Supplementary Table S2. Demographics of the Intervention Cohort.

Supplementary Table S3. Normality test of the phenotypic measurements.

Supplementary Table S4. Estimated heritability for carotenoid species.

Supplementary Table S5. SNPs that are significantly associated with carotenoid metabolism.

Supplementary Table S6. Results of the linear regression test for gene-by-dosage interactions across 37 SNPs.

Supplementary Table S7. Per-base coverage of 35 genes from target sequencing.

Supplementary Table S8. *PKD1L2* variants associated with plasma β-carotene.

Supplementary Table S9. Variants associated with plasma cryptoxanthin, skin carotenoid levels, and β-carotene identified in outlier analysis.

### Supplementary Figures

Supplementary Figure S1. Scree plot of the MDS eigenvalues for the Primary Study Cohort.

Supplementary Figure S2. Density plots of plasma carotenoid subspecies concentrations.

Supplementary Figure S3. Density plots of plasma carotenoid and subspecies concentrations (and skin carotenoids in the East Asian (EAS) and South Asian (SAS) groups.

Supplementary Figure S4. Genome-wide association analysis of skin carotenoid level.

Supplementary Figure S5. Gene-by-dosage plots of plasma carotenoids, genotype, and intervention dosage at week 6.

Supplementary Figure S6. Distribution and effects of genetic variants across selected genes.

Supplementary Figure S7. LocusZoom plot of variants in the *PKD1L2* gene on chromosome 16.

### Supplementary Methods

DNA processing and genotyping

Genotyping data imputation

SNP and individual quality control

Rare Variants Annotation

