## Supplementary material for "Common and rare genetic variation intersects with ancestry to influence human skin and plasma carotenoid concentrations": Carotenoid Genetics Supplementary Information

**(Han et. al)**

### Supplementary Tables

#### Common and rare genetic variation intersects ancestry to influence skin and plasma carotenoid concentrations

#### (Han et. al)

Supplementary Table S1. Demographics of the Primary Study Cohort.

Supplementary Table S2. Demographics of the Intervention Cohort.

Supplementary Table S3. Normality test of the phenotypic measurements.

Supplementary Table S4. Estimated heritability for carotenoid species.

Supplementary Table S5. SNPs that are significantly associated with carotenoid metabolism.

Supplementary Table S6. Results of the linear regression test for gene-by-dosage interactions across 37 SNPs.

Supplementary Table S7. Per-base coverage of 35 genes from target sequencing.

Supplementary Table S8. *PKD1L2* variants associated with plasma β-carotene.

Supplementary Table S9. Variants associated with plasma cryptoxanthin, skin carotenoid levels, and β-carotene identified in outlier analysis.

**Supplementary Table S1. Demographics of the study cohort.**

| Variable | Overall<br>N = 213 | Site 1<br>N = 121[1] | Site 2<br>N = 92[1] | p-value[2] |
|---|---|---|---|---|
| **Ethnic group** | | | | 0.3 |
| Non-Hispanic Black | 61 (29%) | 40 (33%) | 21 (23%) | |
| Asian | 53 (25%) | 25 (21%) | 28 (30%) | |
| Non-Hispanic White | 70 (33%) | 40 (33%) | 30 (33%) | |
| Hispanic | 29 (14%) | 16 (13%) | 13 (14%) | |
| **Sex** | | | | 0.068 |
| Male | 62 (29%) | 29 (24%) | 33 (36%) | |
| Female | 151 (71%) | 92 (76%) | 59 (64%) | |
| **Age (Years)** | 30 (24, 41) | 29 (22, 38) | 31 (25, 46) | 0.032 |

[1]n (%);Participants count from clinic site: North Carolina (1), Minnesota (2)

[2]Fisher's exact test; Wilcoxon rank sum test

**Supplementary Table S2. Demographics of the intervention cohort.**

| Variable | Overall N = 162[1] | Site 1 N = 61[1] | Site 2 N = 41[1] | Site 3 N = 60[1] | p-value[2] |
|---|---|---|---|---|---|
| **Ethnic group** | | | | | 0.36 |
| Non-Hispanic Black | 41 (25%) | 15 (25%) | 12 (29%) | 15 (25%) | |
| Asian | 40 (25%) | 15 (25%) | 10 (24%) | 15 (25%) | |
| Non-Hispanic White | 44 (27%) | 19 (31%) | 10 (24%) | 15 (25%) | |
| Hispanic | 37 (23%) | 12 (20%) | 10 (24%) | 15 (25%) | |
| **Sex** | | | | | 0.044 |
| Male | 79 (49%) | 29 (24%) | 20 (36%) | 30 (50%) | |
| Female | 83 (51%) | 32 (76%) | 21 (64%) | 30 (50%) | |
| **Age (Years)** | 29 (24, 38) | 28 (22, 38) | 28 (25, 25) | 31 (27, 39) | 0.032 |

[1]n (%); Participants count from clinic site: North Carolina (1), Minnesota (2), and Texas (3)

[2]Fisher's exact test; Wilcoxon rank sum test

**Supplementary Table S3. Normality test of the carotenoids' measurements.**

| Measurements | | Original value | | $Log_2$-transformed value | |
|---|---|---|---|---|---|
| **Outcome Category** | **Carotenoid Outcome** | **W** | **p-value** | **W** | **p-value** |
| Plasma | Carotenoid | 0.856 | 4.84E-13 | 0.981 | 6.91E-03 |
| | α-Carotene | 0.671 | 2.2E-16 | 0.935 | 5.40E-08 |
| | β-Carotene | 0.715 | 2.2E-16 | 0.973 | 5.78E-04 |
| | Lycopene | 0.961 | 1.67E-05 | 0.985 | 2.64E-02 |
| | Cryptoxanthin | 0.761 | 2.2E-16 | 0.979 | 3.87E-03 |
| | Lutein/Zeaxanthin | 0.935 | 6.03E-08 | 0.996 | 8.93E-01 |
| Skin | Carotenoid | 0.972 | 3.99E-04 | 0.965 | 4.78E-05 |
| Food | Carotenoid | 0.794 | 9.69E-16 | 0.993 | 4.22E-01 |
| | α-Carotene | 0.545 | 2.2E-16 | 0.969 | 1.89E-04 |
| | β-Carotene | 0.694 | 2.2E-16 | 0.992 | 3.75E-01 |
| | Cryptoxanthin | 0.589 | 2.2E-16 | 0.993 | 5.38E-01 |
| | Lycopene | 0.844 | 1.44E-13 | 0.988 | 8.40E-02 |
| | Lutein/Zeaxanthin | 0.758 | 2.2E-16 | 0.991 | 1.98E-01 |

**Supplementary Table S4. Estimated heritability for carotenoid species.**

| Phenotypes | V(G)/Vp | SE |
|---|---|---|
| Plasma Total Carotenoids | 0.082 | 0.157 |
| Plasma α-Carotene | 0.351 | 0.338 |
| Plasma β-Carotene | 0.157 | 0.203 |
| Plasma Cryptoxanthin | 0.438 | 0.322 |
| Plasma Lutein/Zeaxanthin | 0.091 | 0.172 |
| Plasma Lycopene | 0.168 | 0.211 |
| Skin Carotenoid | 0.301 | 0.302 |

**Supplementary Table S5. SNPs that are significantly associated with carotenoid metabolism.** SNPs were identified using GEMMA and selected with a p-value threshold of < $5\times10^{-8}$ and lambda < 1.04. "Study Cohort by GEMMA" refers to the linear regression analysis on 207 individuals, including covariates for age, sex, BMI, the first two dimensions of the MDS, and dietary carotenoid intake. "Intervention Only Cohort by GEMMA and METAL" refers to the linear regression analysis performed on 110 individuals exclusively from the intervention cohort, using baseline carotenoid measurements as the phenotypic data and the same covariates as in the primary study cohort analysis. These SNPs were further validated through METALysis in the intervention cohort, with corresponding p-values and effect directions reported. "Microarray Genotyped (G) / Imputed (I)" indicates whether SNPs were directly genotyped (G) using the Infinium™ H3Africa Consortium Array v2 or imputed (I) via the Michigan Imputation Server, based on the 1000G Phase 3 v5 (GRCh37/hg19) reference panel. The "Nearest Gene" was identified using SNPnexus web tools.

Excel table

**Supplementary Table S6. Results of linear regression tests for gene-by-dosage across 37 SNPs.** These SNPs were identified based on significant associations within the study cohort and further supported by signals in the intervention cohorts.

Excel table

**Supplementary Table S7. Per-base coverage of 35 genes from target sequencing.** These genes were reported to be relevant to carotenoid metabolism and was manually curated from the literature.

| Gene Symbol | GRCh38 coordinates | Average median coverage per base | Proportion of bases with <10X coverage |
|---|---|---|---|
| *ABCA1* | chr9:104784274-104903822 | 1850.61 | 0.029 |
| *ABCB1* | chr7:87504126-87600270 | 1969.27 | 0 |
| *ABCG2* | chr4:88092126-88140057 | 2008.08 | 0 |
| *ABCG5* | chr2:43813056-43838864 | 1637.05 | 0.007 |
| *ABCG8* | chr2:43838987-43878093 | 1713.18 | 0 |
| *APOA1* | chr11:116835747-116837510 | 1192.08 | 0 |
| *APOB* | chr2:21001695-21044021 | 1820.98 | 0 |
| *BCO1* | chr16:81238796-81290655 | 1720.79 | 0 |
| *BCO2* | chr11:112175542-112217939 | 2211.23 | 0.007 |
| *CD36* | chr7:80646713-80674245 | 1659.81 | 0 |
| *CETP* | chr16:56961940-56983766 | 1742.2 | 0.038 |
| *CLPS* | chr6:35795044-35797345 | 1868.47 | 0 |
| *COBLL1* | chr2:164685901-164842263 | 1754.94 | 0.033 |
| *CXCL8* | chr4:73740583-73742530 | 2350.26 | 0 |
| *CYP26B1* | chr2:72132180-72147954 | 1423.63 | 0.010 |
| *ELOVL2* | chr6:10983620-11044278 | 1730.4 | 0.036 |
| *GSTP1* | chr11:67583796-67586615 | 1388.76 | 0 |
| *INSIG2* | chr2:118096502-118108430 | 1797.29 | 0 |
| *IRS1* | chr2:226794977-226798778 | 1816.24 | 0.027 |
| *ISX* | chr22:35066958-35085846 | 1927.92 | 0 |

| | | | |
|---|---|---|---|
| *LDLR* | chr19:11089514-11131434 | 1980.62 | 0 |
| *LIPC* | chr15:58431962-58568878 | 1527.38 | 0 |
| *LPL* | chr8:19939399-19965351 | 1903.95 | 0 |
| *MC4R* | chr18:60371297-60372380 | 1551.05 | 0 |
| *MTTP* | chr4:99564048-99622901 | 2151.1 | 0.022 |
| *NPC1L1* | chr7:44513401-44541311 | 1500.75 | 0.038 |
| *PKD1L2* | chr16:81101012-81220480 | 1951.87 | 0.030 |
| *PNLIP* | chr10:116546025-116567972 | 1913.42 | 0 |
| *RPE65* | chr1:68429744-68450057 | 1854.69 | 0 |
| *SCARB1* | chr12:124778364-124863772 | 1381.6 | 0 |
| *SETD7* | chr4:139496237-139556288 | 2103.21 | 0 |
| *SLC27A6* | chr5:128966025-129033308 | 1968.51 | 0.040 |
| *SOD2* | chr6:159682452-159693231 | 1265.13 | 0 |
| *STARD3* | chr17:39653465-39662949 | 1385.05 | 0 |
| *TCF7L2* | chr10:112950725-113166097 | 1178.52 | 0.021 |

**Supplementary Table S8.** ***PKD1L2*** **variants associated with plasma β-carotene.**

| rsID | P value[*] | P value[†] | P value[‡] | P value[§] | Study MAF | S | F | P2 | P | L | MAF | RefAA | AltAA |
|---|---|---|---|---|---|---|---|---|---|---|---|---|---|
| rs79139155 | 0.0007 | 0.0004 | | | 0.0048 | . | D | D | N | N | 0.0059 | L | F |
| rs4889261 | 0.0019 | 0.0081 | 0.1261 | 0.0284 | 0.8167 | . | N | . | N | N | 0.8098 | L | P |
| rs539835443 | 0.0022 | 0.0017 | . | . | 0.0024 | . | N | . | . | . | 0.0004 | R | T |
| . | 0.0022 | 0.0017 | | | 0.0024 | . | . | . | . | . | . | . | . |
| rs370822268 | 0.0022 | 0.0017 | 0.5565 | 0.8277 | 0.0024 | . | D | . | . | . | 0.0004 | E | G |
| rs537876996 | 0.0022 | 0.0017 | 0.5565 | 0.8277 | 0.0024 | . | . | . | . | . | . | . | . |
| . | 0.0022 | 0.0017 | | | 0.0024 | . | N | . | N | . | 0.0000 | V | I |
| rs187577452 | 0.0022 | 0.0017 | | | 0.0024 | . | . | . | . | . | . | . | . |
| rs7194871 | 0.0034 | 0.0123 | | | 0.8024 | . | N | . | N | N | 0.7937 | K | Q |

[*]Race/ethnicity included as a covariate

[†]Mutiscaling Dimentions included as covariate

[‡]Intervention Cohort (Race/ethnicity included as a covariate)

[§]Intervention Cohort (No covariate)

S; SIFT, F; fathmm, P2; Polyphen2, P; ROVEAN, L; LRT, RefAA; Reference Amino Acid, MAF; gnomAD_genomes_AF, AltAA; Alternate Amino Acid

**Supplementary Table S9. Variants associated with plasma cryptoxanthin, skin carotenoid levels, and β-carotene identified in outlier analysis.**

| rsID | Gene | β | p-value | Study MAF | No. | S | F | P2 | P | L | MAF | RefAA | AltAA |
|---|---|---|---|---|---|---|---|---|---|---|---|---|---|
| rs2303790 | *CETP* | 1.3388 | 0.0152 | 0.0048 | 2 | D | D | . | D | U | 0.0015 | D | G |
| rs756535387 | *APOA1* | 2.2842 | 0.0031 | 0.0024 | 1 | D | D | D | D | U | . | R | C |
| rs142824860 | *BCO1* | -3.7005 | 0.0013 | 0.0047 | 1 | D | D | D | D | D | 9.72E-05 | E | G |

S; SIFT, F; fathmm, P2; Polyphen2, P; ROVEAN, L; LRT, RefAA; Reference Amino Acid, MAF; gnomAD_genomes_AF, AltAA; Alternate Amino Acid

### Supplementary Figures

#### Common and rare genetic variation intersects with ancestry to influence human skin and plasma carotenoid concentrations

#### (Han et. al)

Supplementary Figure S1. Scree plot of the MDS eigenvalues for the Primary Study Cohort.

Supplementary Figure S2. Density plots of plasma carotenoid species concentrations.

Supplementary Figure S3. Density plots of plasma carotenoid and species concentrations (and skin carotenoids in the East Asian (EAS) and South Asian (SAS) groups.

Supplementary Figure S4. Genome-wide association analysis of skin carotenoid level.

Supplementary Figure S5. Gene-by-dosage plots of plasma carotenoids, genotype, and intervention dosage at week 6.

Supplementary Figure S6. Distribution and effects of genetic variants across selected genes.

Supplementary Figure S7. LocusZoom plot of variants in the *PKD1L2* gene on chromosome 16.

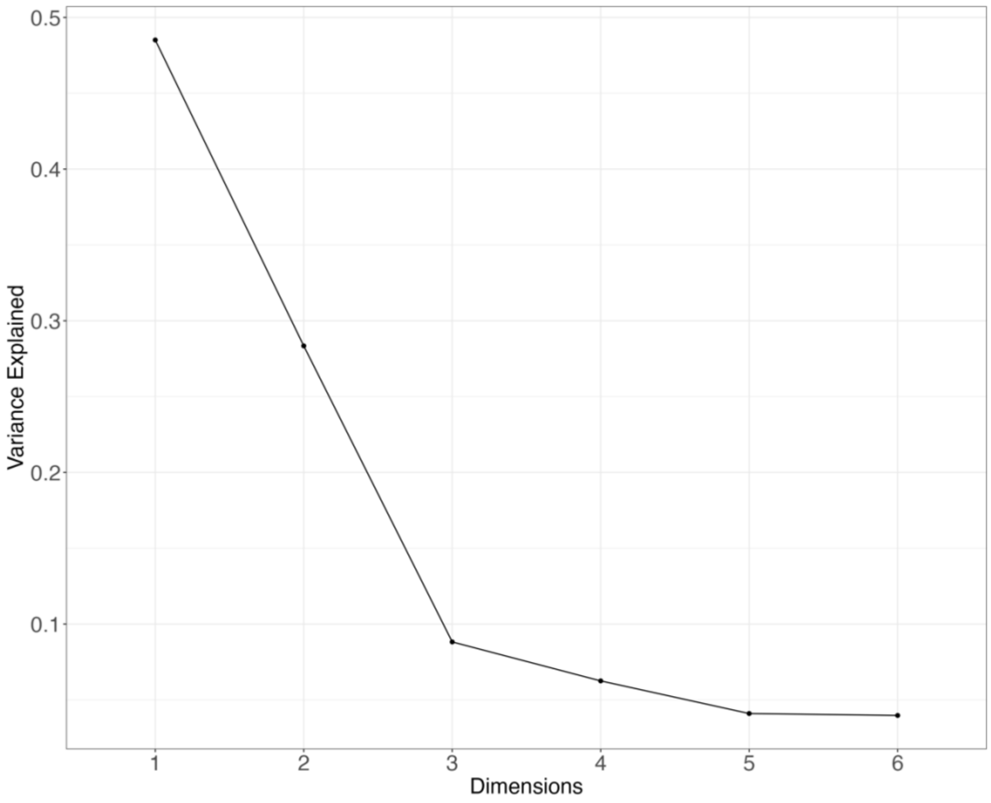


**Supplementary Figure S1. Scree plot of the MDS eigenvalues for the Primary Study Cohort.** The first two dimensions explain 48.5% and 28.3% of the total variance, respectively, accounting for 76.8% of the ancestry-related variation in the data.

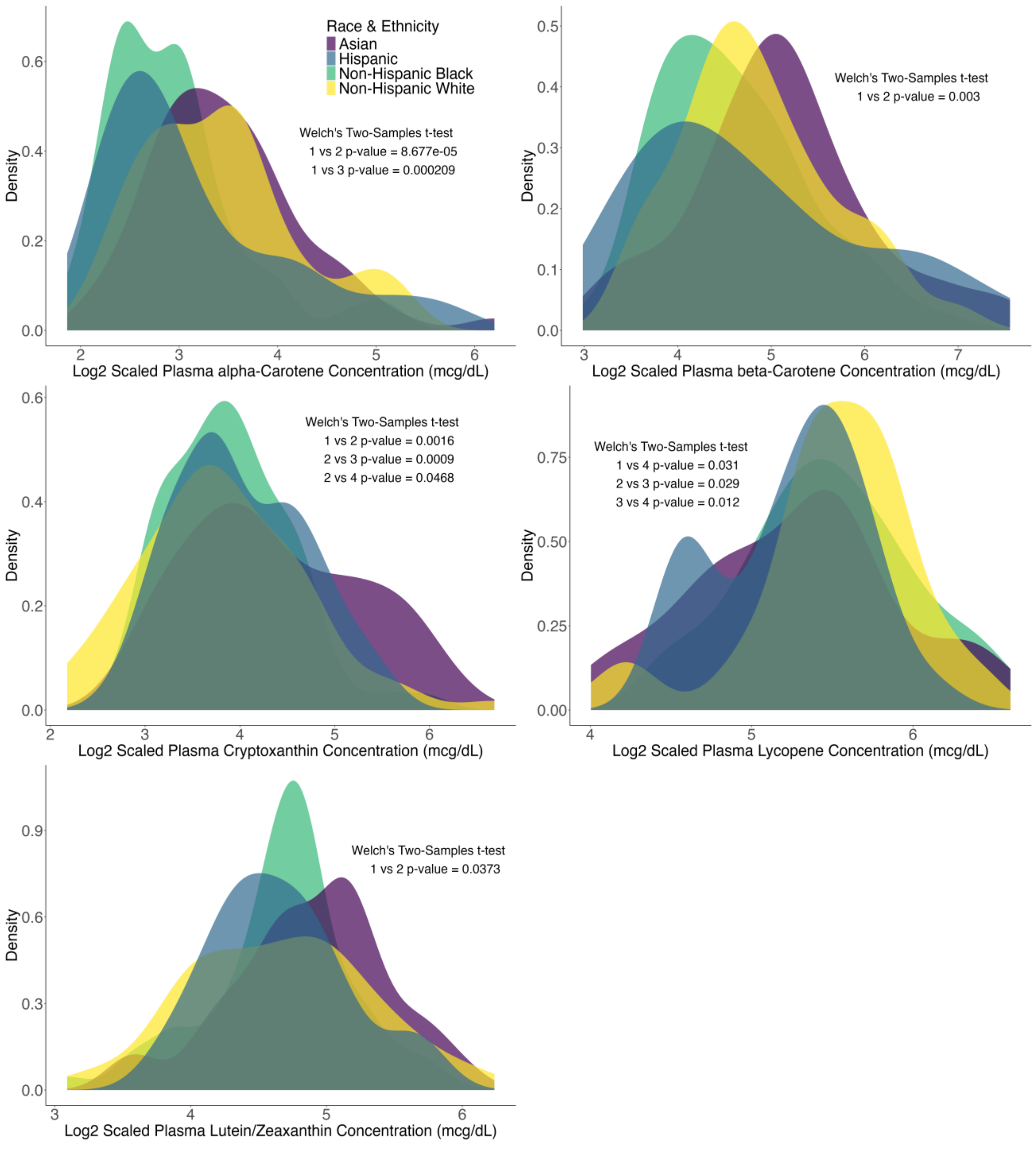


**Supplementary Figure S2. Density plots of plasma carotenoid species concentrations.** Non-Hispanic Black (1), Asian (2), Non-Hispanic White (3), and Hispanic (4). Welch's Two-Samples t-test reveals significant variation in plasma carotenoid metabolism between these groups, suggesting racial and ethnic influences on carotenoid concentration.

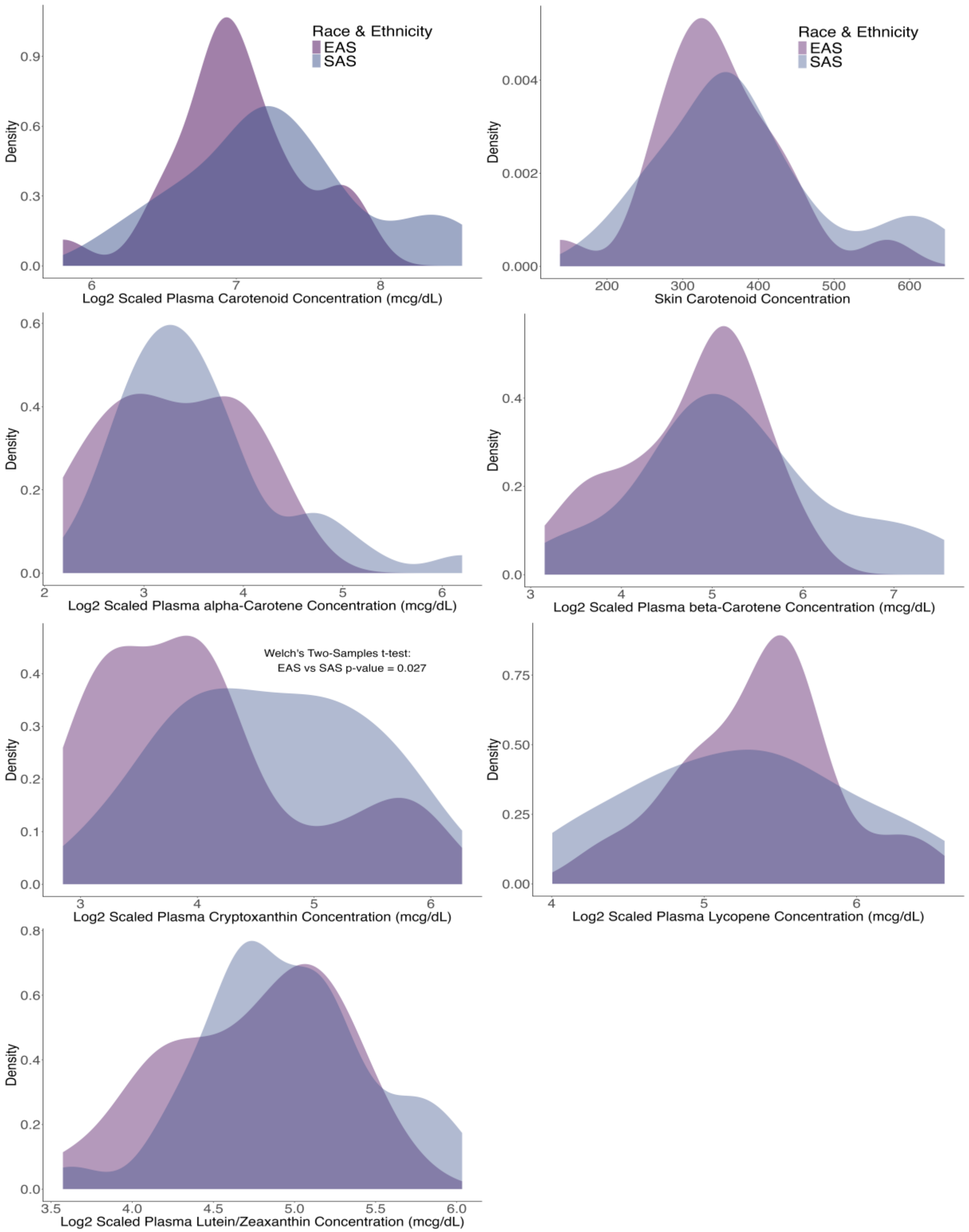


**Supplementary Figure S3. Density plots of plasma carotenoid and species concentrations (and skin carotenoids in the East Asian (EAS) and South Asian (SAS) groups.** Welch's Two-

Samples t-test indicates no significant differences in carotenoid metabolism between the two groups.

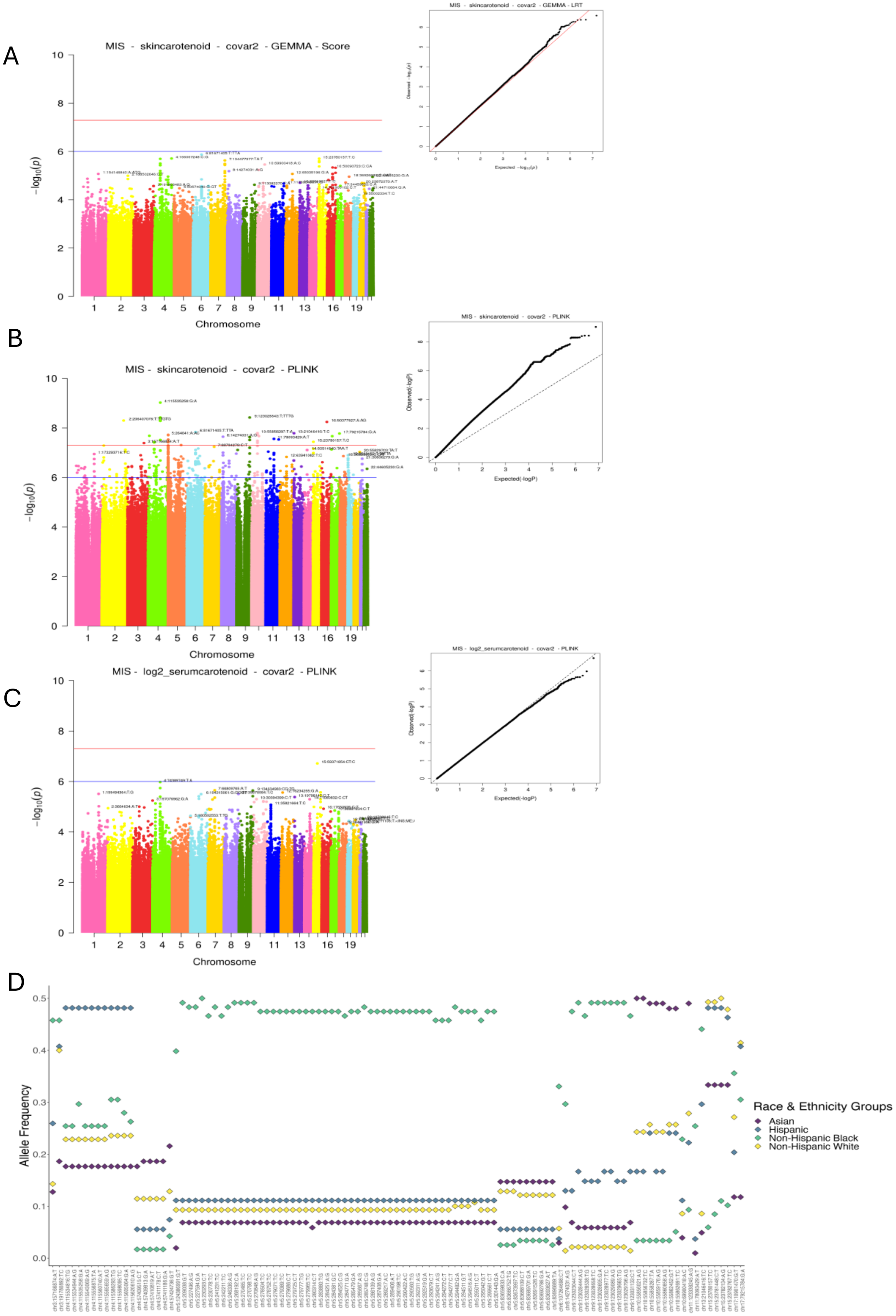
A
MIS - skincarotenoid - covar2 - GEMMA - Score
MIS - skincarotenoid - covar2 - GEMMA - LRT
B
MIS - skincarotenoid - covar2 - PLINK
MIS - skincarotenoid - covar2 - PLINK
C
MIS - log2_serumcarotenoid - covar2 - PLINK
MIS - log2_serumcarotenoid - covar2 - PLINK
Chromosome
Expected(-logP)
Observed(-logP)
D
Allele Frequency
Race & Ethnicity Groups
Asian
Hispanic
Non-Hispanic Black
Non-Hispanic White

**Supplementary Figure S4. Genome-wide association analysis of skin carotenoid level. S4A**. Manhattan and Q-Q plots from GEMMA linear regression analysis of skin carotenoid levels. **S4B** and **S4C**. Manhattan and Q-Q plots from PLINK linear regression results for skin carotenoid (**S4B**) and $\log_2$-transformed total plasma carotenoid concentration (**S4C**). The Manhattan plots highlight loci reaching the genome-wide significance threshold (blue line at -logP = 6) or the SNP significance threshold (red line at -logP = 7.3). The Q-Q plots evaluate the distribution of observed versus expected p-values. **S4D**. Scatter plot shows ancestral minor allele frequency differences across four racial/ethnic groups for SNPs surpassing the genome-wide threshold in (**S4B**), emphasizing how genetic ancestry influences variability in skin carotenoid levels, primarily driven by minor allele frequency differences.

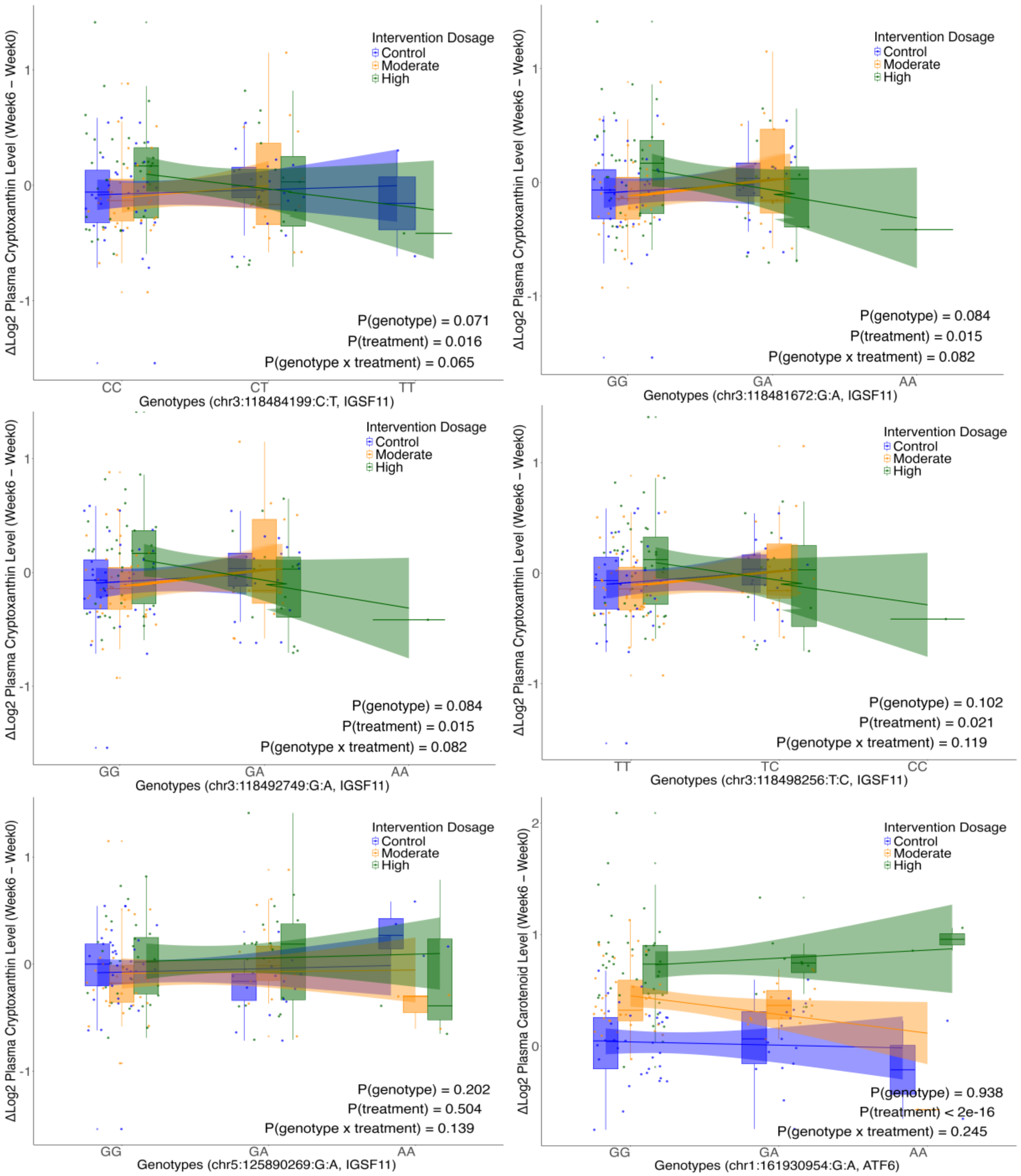


**Supplementary Figure S5. Gene-by-dosage plots of plasma carotenoids, genotype, and intervention dosage at week 6.** The genotypes are categorized along the x-axis, with each intervention dosage (Control, Moderate, High) represented by distinct colors: blue for Control, orange for Moderate, and green for High. Box plots show the distribution of plasma carotenoid levels within each genotype and dosage group, while smoothed regression lines highlight trends within dosage levels. The p-values indicate the significance of the genotype effect (P(genotype)), treatment effect (P(treatment)), and their interaction effect (P (genotype x treatment)). The significant p-value for treatment indicates an overall dosage effect on plasma carotenoid levels, while the genotype-by-treatment interaction p-value suggests a near-significant trend, implying a possible interaction effect.

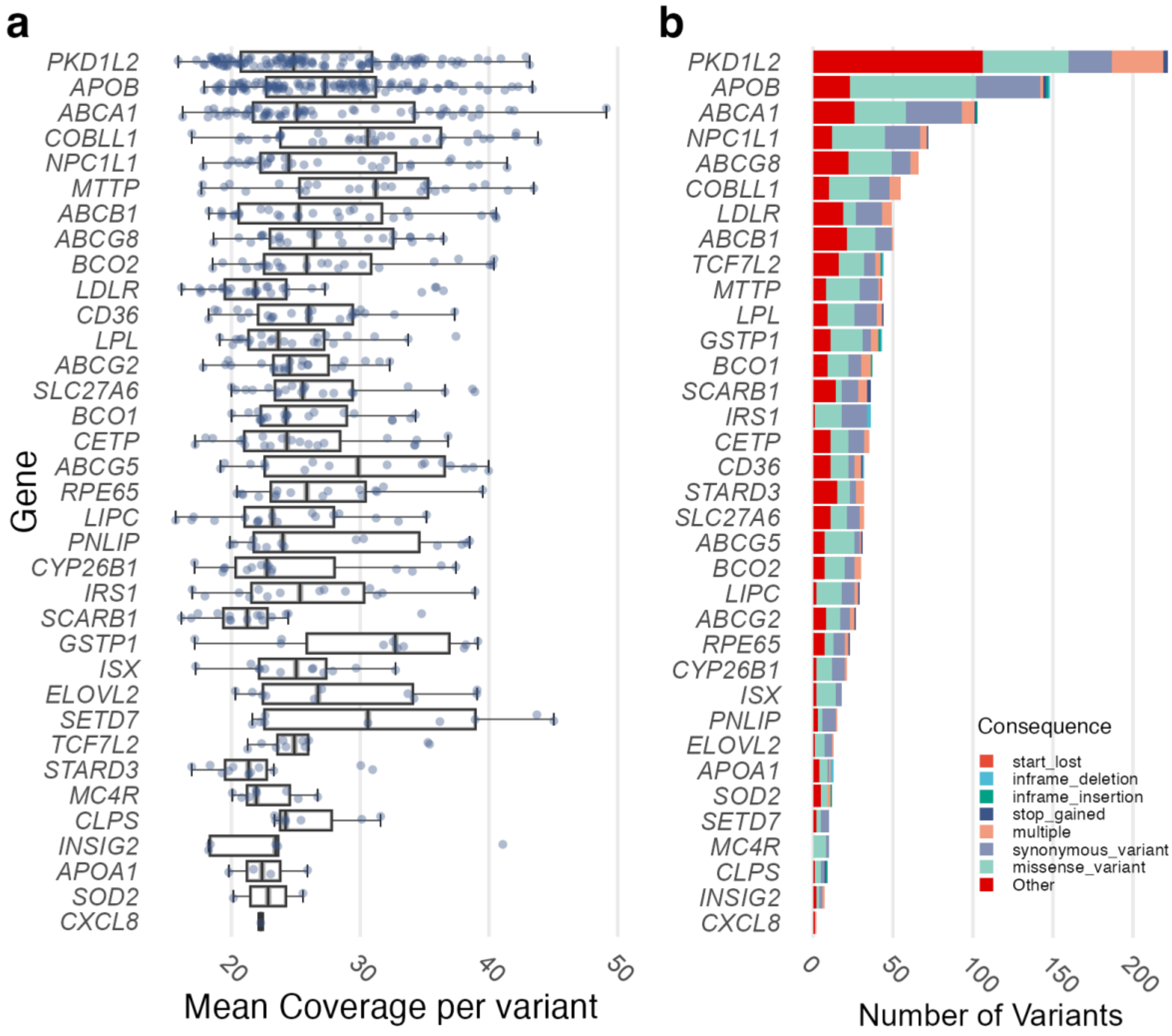


**Supplementary Figure S6. Distribution and effects of genetic variants across selected genes.** **(a)** Box plot displaying the depth of coverage for genetic variants post-quality control (QC) across various genes, with individual points jittered for clarity. Each dot represents a single variant, with the x-axis showing the depth of coverage and the y-axis listing the genes. **(b)** Stacked bar graph illustrating the number of genetic variants classified by their effect types for each gene. The x-axis represents the count of variants, and the y-axis lists the genes. Bars are color-coded to distinguish different variant effects, with the length of each bar segment representing the count of variants with a specific effect within each gene. Larger genes, such as *PKD1L2*, *APOB*, *ABCA1*, and *NPC1L1*, exhibit a greater number and diversity of variant effects, while smaller genes, such as *CXCL8*, *SOD2*, and *INSIG2*, show fewer variants with a more limited range of effect types.

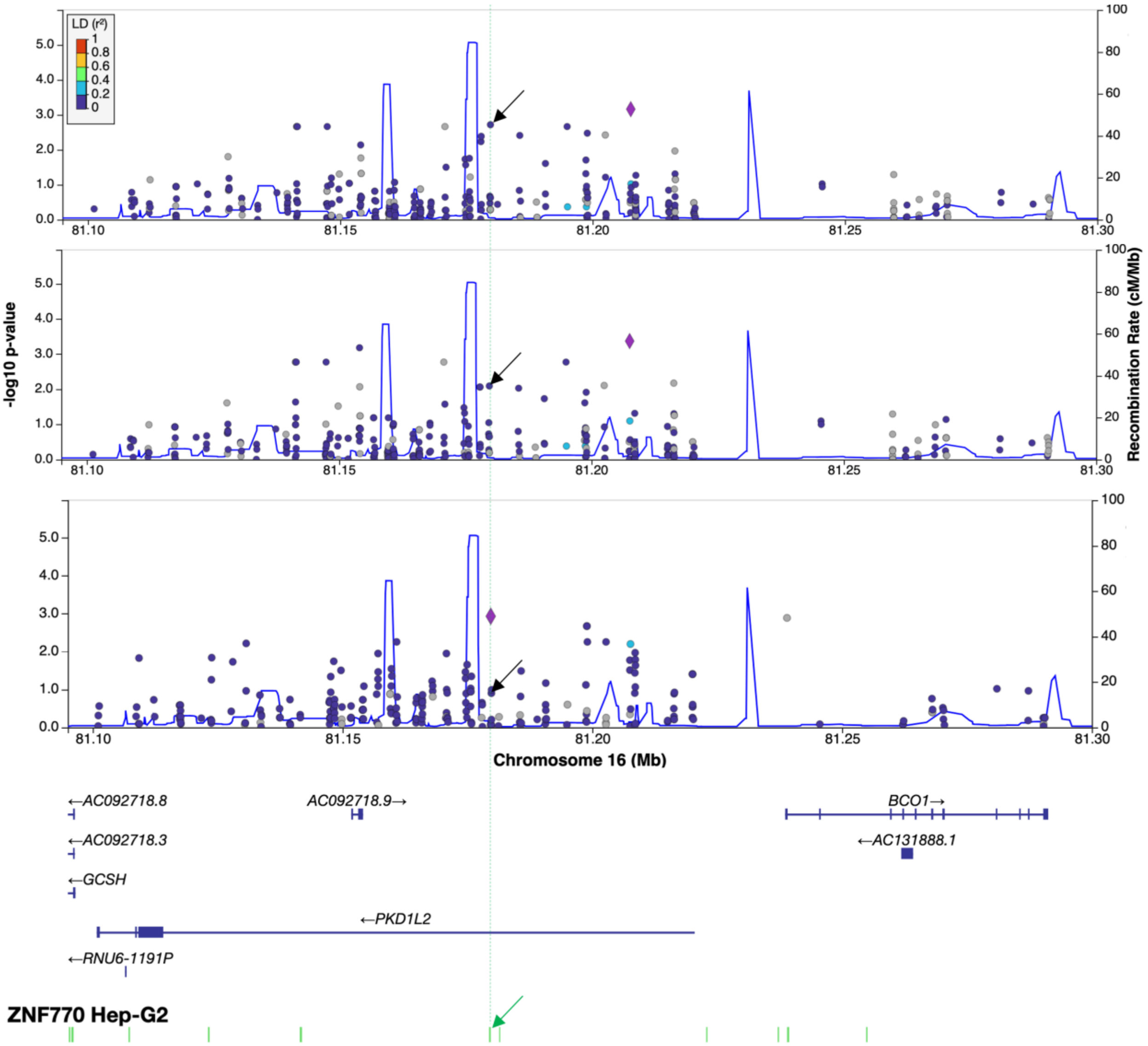


**Supplementary Figure S7. LocusZoom plot of variants in the *PKD1L2* gene on chromosome 16.** These plots illustrate the association of SNVs in the *PKD1L2* gene region on chromosome 16 with plasma β-carotene concentration. The y-axis displays the $-\log_{10}$ p-values and the x-axis displays genomic coordinates (GRCh38). **S7A.** Primary analysis using standard covariates, with rs79139155 as the lead SNV (purple diamond). **S7B**. Analysis replacing MDS dimensions, also highlighting rs79139155. **S7C**. Replication cohort analysis, highlighting rs4889261 the only variant with nominal significance across all analyses (in a model without covariates). Top SNVs are represented by a diamond shape. Point colors indicate LD ($r^2$) with the lead SNV, and the dotted vertical line marks overlap of rs4889261 (black arrows) with ZNF770 ChIP-Seq regulatory sites in Hep-G2 cells (Green arrow).

### Supplementary Methods

#### DNA processing and genotyping

Genomic DNA was isolated from the buffy coat fraction of blood collected at the participant visits, using the Gentra Puregene Kit (Qiagen, Germany). Samples were genotyped on the Infinium™ H3Africa Consortium Array v2 (Illumina, USA). The array was designed by the H3Africa consortium to encompass common variants (minor allele frequency (MAF) $\geq 0.01$) present in at least one of the African populations represented in the H3Africa Consortium sequencing study[1], as well as medically relevant variants from available databases, including ACMG[2], ClinVar[3], COSMIC[4], PharmGKB[5]. The array includes assays for ~2.8 million SNPs. The genotyped dataset was initially uploaded to GenomeStudio v2.0.5, where non-performing SNPs (>4 SD out of HWE, or missing in >10% SNPs with visual evidence of skewed clustering) were removed. A final PLINK[6]-compatible file containing 2,271,503 SNPs was generated using the PLINK Exporter plugin, which was then used for downstream QC and analysis.

#### Genotyping data imputation

To generate valid VCF files before phasing, imputation, and association tests, we corrected for monomorphic sites, ensured consistency of reference alleles with the reference genome, addressed variants with invalid genotypes, and removed monomorphic sites. The filtered SNPs were then verified using the checkVCF.py Python script (https://github.com/zhanxw/checkVCF). The genotype data was phased and imputed using EAGLE 2.4[7]. The quality-controlled genotyping dataset, comprising a total of 1,917,156 variants, was then imputed using the 1000G Phase 3 v5 (GRCh37/hg19) reference panel on the Michigan Imputation Server[8], resulting in 26,084,710 SNPs. These SNPs were filtered for $r^2 \geq 0.3$ and MAF ≥ 0.05. A refined set of 7,467,403 high-quality SNPs was identified for downstream analyses.

##### SNP and individual quality control

To address potential population stratification in our genotyping dataset, we employed PLINK[6] 1.9. The array data were re-filtered with the following parameters to remove low-quality variants: a minor allele frequency (MAF) threshold of 0.05 (--maf 0.05), a per-individual missingness threshold of 0.1 (--mind 0.1), a per-genotype missingness threshold of 0.1 (--geno 0.1), and a deviation from Hardy-Weinberg equilibrium threshold of 0.001 (--hwe 0.001). Pairwise identity-by-descent (IBD) assessment was performed on the resulting pruned dataset, and the pairs with a proportion of IBD (PI_HAT) score greater than 0.25 (PI_HAT > 0.25) were flagged, then the individual within the pair with the higher missingness rate was removed from the cohort. Strand-ambiguous SNPs and those in high linkage disequilibrium were pruned using the --indep-pairwise flag with a window size of 50 SNPs, a sliding window of 10, and $R^2$ values greater than 0.1.

##### Rare Variants Annotation

Damaging variants were identified and annotated using multiple predictive algorithms to assess their functional impact. Variants were first filtered for ultra-rare frequency (minor allele frequency [MAF] < 1.0E-05) using the gnomAD database. Functional predictions were obtained from SIFT[9], fathmm[10], Polyphen2[11], and PROVEAN[12], with each tool categorizing variants as deleterious based on their specific scoring systems. Additionally, pathogenicity was evaluated using AlphaMissense and CADD[13], providing a probability score for likely pathogenicity. Variants with combined evidence of deleterious impact across these platforms were considered high-impact candidates.
